# *Plasmodium falciparum* Carriage in central and northern Mali amid Seasonal Malaria Chemoprevention implementation

**DOI:** 10.1101/2025.11.26.25341058

**Authors:** Betty Kazanga, Mady Cissoko, Abdoulaye Katile, El Bahdja Boudoua, Eva Legendre, Bourama Kamate, Moussa Bamba Kanoute, Paul Kone, Abdoulaye Djiguiba, Ousmane dit Abel Poudiougo, Moulaye Coulibaly, Amatigue Zeguime, Antoine Claessens, Coralie L’Ollivier, Maissane Mehadji, Stéphane Ranque, Marc-Karim Bendiane, El-Hadj Konco Ciré Ba, Abdoulaye Djimde, Jean Gaudart, Issaka Sagara, Jordi Landier

**Affiliations:** Aix Marseille Univ, IRD, INSERM, SESSTIM, ISSPAM, Marseille, France; National malaria control program, Ministry of Health and Social Development, Bamako, Mali; Malaria Research and Training Center, University of sciences, Techniques and Technologies of Bamako, Bamako, Mali; Hôpital Européen, Marseille France.; District hospital (CSREF) of Dire, Region of Tombouctou, Mali; LPHI, CNRS, INSERM, University of Montpellier, Montpellier, France.; Aix-Marseille Univ., AP-HM, SSA, RITMES, 13005 Marseille, France.; IHU Méditerranée Infection, 13005 Marseille, France.; Service de médecine interne, gériatrie et thérapeutique, Assistance publique hôpitaux de Marseille (AP-HM), Marseille, France; ADES UMR 7268, AMU, CNRS, EFS, Marseille, France; Aix Marseille Univ, IRD, MINES, IRD-UCAD International Campus, Dakar, Senegal.; Aix Marseille Univ, IRD, INSERM, SESSTIM, ISSPAM, AP-HM, Hop. La Timone, BioSTIC, Marseille, France.

**Keywords:** Malaria, *Plasmodium falciparum*, Asymptomatic carriage, Seasonal Malaria Chemoprevention, persistence

## Abstract

**Introduction:** Seasonal malaria chemoprevention (SMC) reduces malaria incidence in children under five in Sahelian countries. However, transmission persists at substantial levels and incidence is re-increasing. Addressing *Plasmodium falciparum* carriage in asymptomatic individuals could represent a relevant intervention to complement current access to care- and vector-control-based strategies. In this study, we aim to characterise the parasite reservoir dynamics over one year in the general population of two sites of central and northern Mali.

**Methods:** We included members of randomly selected households in four villages (2 in Kati and 2 in Dire districts). At four planned visits, infection was detected using rapid diagnostic tests (RDTs) and qPCR. Clinical malaria incidence was recorded passively at local health facility and *Plasmodium* positive samples were genotyped. We analyse clinical malaria incidence, *P. falciparum* PCR prevalence, parasite density and multiplicity of infection (eMOI) at different seasons, as well as factors associated with *P. falciparum* carriage.

**Results:** Malaria seasonal dynamics was driven by rainfall in Kati (Central Mali) and by flooding in Dire (Northern Mali). In Kati, prevalence rose from 21.5% at the dry season 2021 baseline to 33.5% in December after the rainy season. Prevalence was lowest in SMC-eligible children across 3 of 4 surveys. Prevalence was highest (40-50%) for 10-24 years old in December. Median parasite densities were generally <10 parasites/µL except for 10-24 years old, who also presented higher eMOI. In Dire, prevalence was high at both baseline and December survey (60%) but dropped in September and in May 22. SMC was not distributed in 2021, and prevalence was homogenously high (50-70%) across groups in December. Parasite densities and eMOI fluctuated strongly across age-groups and survey.

**Discussion:** Our findings show that in the “classic” Sahelian, rainfall-driven setting of Kati, a high prevalence of asymptomatic infections among 10-24 year-olds likely represents the reservoir for transmission. In the understudied Northern Mali, our results suggest generalized high burden and complex dynamics. Continuous SMC modifies the age-distribution of the parasite reservoir. Targeting asymptomatic carriers beyond SMC coverage could represent a relevant strategy to further reduce transmission in seasonal settings.

## Introduction

In 2023, Mali remained among the 11 nations with the highest *Plasmodium falciparum* malaria burden, reporting an estimated 8.2 million cases (3.1% global cases) and 14,200 deaths (2.4% global deaths) [1]. Clinical malaria incidence peaks during the rainy season (July-November), prompting intensified control efforts including Seasonal Malaria Chemoprevention (SMC).

Since 2012, SMC in Mali consists of monthly administration of sulfadoxine-pyrimethamine plus amodiaquine (SP-AQ) to children aged 3–59 months during the rainy season with 3 to 5 cycles annually depending on the local transmission intensity. In 2021, coverage reached 83-86% per cycle; 79% completed four cycles meeting coverage targets [2,3].

SMC protects for 28 days reducing clinical malaria incidence by 75%–90% and parasitemia by 62% in targeted children [4,5]. Yet, its broader impact on transmission remains poorly characterised. A 2021 Gambian study showed that increased SMC coverage was associated with a reduced risk of infection for non-eligible members of the same household [6]. This study was however conducted during the year when SMC-eligibility age was extended from 59 months to 10 years old, which could represent a transient higher benefit situation in which those immediately above the new SMC-eligibility age have been exposed to full transmission for 5 years. However, Eastern Senegal “red zone” regions of Kedougou, Kolda and Tambacounda demonstrate that SMC up to 10 years, in combination with vector control and improved access to early diagnostic and treatment for over 10 years, does not prevent is unable to reduce transmission below certain levels [5,7].

Current control interventions are not designed to address chronic *P. falciparum* infections, which generally present as subclinical or even asymptomatic, and often at low parasite densities. Asymptomatic *P. falciparum* infections maintain dry season persistence and transmission restoration when conditions become favourable [8,9]. Pre-SMC studies in the Sahel show that while children under 5 years carried the heaviest clinical burden, children under 5 or aged 5-15 years carried most asymptomatic infections and represented the largest contributors to the asymptomatic reservoir[10,11], with cumulative exposure likely explaining the difference between settings[12].

Recent studies performed in the SMC context show a lower prevalence in SMC-eligible groups, but persistence of infections in SMC-ineligible age-groups [7,13]. Even with SMC protecting children up to 10 years, persistent reservoirs sustain the parasite pool between seasons and hinder elimination. Their persistence is shaped by naturally acquired immunity, developed through repeated exposure, which modulates parasite density, clinical presentation, and parasite gene expression patterns such as *var* gene profiles [9,14].

In this study, we aim to extend the characterization of the structure and seasonal dynamics of the reservoir in Sahelian settings under routine SMC implementation. We focused on human hosts and parasite population in two distinct epidemiological settings, one with classic “Sahelian” features in Central Mali, and one in the understudied region of Niger delta, in Northern Mali.

## Methods

### Study design and setting

We conducted a prospective cohort study in four villages of Central and Northern Mali. In Central Mali, the study was carried out in villages of Safo (3977 inhabitants) and Torodo (1293 inhabitants)located in Kati health district, approximately 15km northwest of Bamako, where malaria transmission is highly seasonal, peaking between June and December [15,16]. In Northern Mali, the study took place in the Dire health district, in Bourem Sidi Amar (3422 inhabitants) and Koigour (1331 inhabitants) villages, located in the Niger River inner delta, which floods from August to February. This results in a bimodal malaria transmission pattern, with peaks in August–September and again in December–February/March [17]. In 2021, SMC (Four rounds) ran in Kati but was cancelled in Dire due to SMC drug shortages. Kati’s cohort was closed; Dire had new participants at the first follow-up survey (T1).

### Participant enrollment and ethics

A census was conducted in all four villages. Households were randomly selected and heads of selected household were approached for verbal consent to participate. All members present at baseline were enrolled after written informed consent obtained for participants aged 18 and older or from a parent or legal guardian for participants <18 years. Participants aged 12-17 years provided a written assent. Participants who were unable to read or write were assisted by a literate witness. The study protocol was approved by the Ethics Committee of the University of Sciences, Techniques and Technologies of Bamako (protocol No. 2020/297/CE/FMOS/ FAPH).

### Follow-up and data collection

#### Survey visits

Four surveys were conducted to detect asymptomatic *P. falciparum* infections and administer structured questionnaires: T0 during the dry, low transmission season; T1 immediately before the first round of SMC; T2 at the end of the wet season; T3 during the next dry season. In Kati, T0 took place mid-May; T1 mid-July (starting on 15 July, as SMC round 1); T2 early December and T3 mid-May. In Dire, T0 took place in June 2021, T1 at the end of August, T2 mid-December, and T3 at the end of May 2022. During visits, tablet-based questionnaires captured sociodemographic information, factors potentially associated with *P. falciparum* infection (bed net use, infection, treatment, nighttime outdoor activities, and travel). Finger-prick blood was tested using a rapid diagnostic test (RDT; SD Bioline Malaria Ag Pf, Abbott Diagnostics Korea, Gyeonggi-do, South Korea), a thick smear for microscopy and a dried blood spot for polymerase chain reaction (PCR). RDT-positive participants in Dire were treated regardless of symptoms; in Kati only symptomatic participants were treated according to national malaria treatment guidelines.

#### Follow-up between surveys

Between surveys, weekly households were conducted in Dire to record participant mobility (mainly presence, absence, and date of departure). In Bourem Sidi Amar (Dire), Torodo and Safo (Kati), clinical cases were passively recorded at health facilities located in each village. In Koigour (Dire), participants had to reach Bourem Sidi Amar health post (5km distance). Clinical registries were then digitized using REDCap.

#### Laboratory analyses

*P. falciparum* infection was detected and parasite density (parasites per microliter of blood) estimated by quantitative PCR (qPCR) performed in duplicate. One assay targeted the multicopy *P. falciparum* var acidic terminal sequence (*varATS*) gene and the other the *CytB* gene. A result was considered positive for a gene (varATS or CytB) if both duplicates were positive or ≥2 out of 4 in case of initially discordant result repeated. A sample was considered positive if it was positive for at least one gene (varATS and of CytB). Detailed methods, including sample processing, DNA extraction, and assay conditions, were described previously[7].

PCR-positive samples were genotyped by SpotMalaria consortium at Wellcome Sanger Institute using amplicon-based sequencing method[18]. The output consisted of 101 bi-allelic SNPs located on the 14 chromosomes and concatenated into a ‘molecular barcode’, plus six markers of resistance to antimalarials (*aat1* S528L, *crt* K76T, *dhfr* S108N, *dhps* A437G, *kelch13* C580Y and *mdr1* N86Y). Genetic barcode SNPs had been picked for their variable allele frequencies within the *P*. *falciparum* population.

#### Data analysis and statistical methods

All analysis was conducted using R software version 4.5.1 [19] and stratified by site.

#### Malaria incidence

In this study, clinical cases referred to RDT-confirmed infections passively detected by health facilities between visits, and asymptomatic *P. falciparum* infections referred to qPCR-positive infections detected at survey visits.

The cumulative incidence between survey visits was calculated as the number of clinical cases divided by the total person-time at risk. In Dire, weekly incidence was calculated by dividing the number of RDT-confirmed cases each week by the number of participants under follow-up that week. In Kati, where weekly mobility data were unavailable, follow-up time was estimated based on survey visit attendance: 100% for participants who were present at two consecutive visits, 50% for those present at one visit, and no follow-up time for those not present at any survey visit.

#### Malaria prevalence

*P. falciparum* prevalence was defined as the proportion of qPCR-positive individuals at a survey visit. Prevalence by visit and age group was calculated as the number of positives divided by the total tested in each group. To estimate 95% confidence intervals of prevalence estimates, we used the {survey} package in R, accounting for a simple cluster sampling design with varying cluster sizes [20,21].

#### Parasitemia analysis

We evaluated parasite density patterns among infected individuals and assessed RDT sensitivity in the cohort. Parasitemia was categorised into five ranges: <3 (positive below the quantification limit of the assay), 3–10, 10–100, 100–1000, and >1000 parasites/µL. For each age group and survey, we calculated the proportion of samples within each density range and the corresponding RDT sensitivity.

#### Multiplicity of infection

Multiplicity of Infection (MOI), also known as complexity of infection (COI), is a genetic metric that quantifies the number of distinct *P. falciparum* strains simultaneously infecting a single host. MOI provides insights into the number of distinct parasite lineages within individuals and overall transmission intensity[22,23]. Using the barcodes obtained from MalariaGEN Spot Malaria, the effective multiplicity of infection (eMOI) was estimated with the Bayesian MOIRE R package, which adjusts the conventional MOI to account for relatedness among parasite clones within each infection. This approach accommodates situations in which parasites within a host may be genetically related rather than entirely independent[24].

eMOI is defined as:

eMOI=MOI×(1−r_w_)+1

Where r_w_ is the average genomic relatedness among clones within a host.

Infections with eMOI > 1.1 were considered polyclonal (multiple parasite genotypes). Those with eMOI ≤ 1.1 were classified as monoclonal (single parasite genotype), which is a practical threshold supported by several studies[22,25]. An eMOI of 1.5 can be interpreted as a mixed infection where most parasites share one genotype, whereas an eMOI of 2 could be two unrelated parasite lineages, or three parasite lineages, of which two are related.

#### Factors associated with *Plasmodium falciparum* carriage

We defined *P. falciparum* carriage as qPCR-positive infection at surveys. We analysed factors associated with carriage using generalised additive multilevel logistic regression models (GAMM) in the {mgcv} package[26]. Explanatory variables included age, sex, participation in SMC, correct bed net use (reported daily use without body parts outside the net), nighttime outdoor activities, and mobility. Previous clinical malaria and carriage at the preceding survey visit were also included to account for persistence or recurrence of infections. Nighttime activities and mobility were summarised into profiles using principal component analysis (PCA) followed by hierarchical clustering (HCPC) (Additional File: Methods S1) [27].

Three models were fitted. The first examined age and sex across all four surveys, with random intercepts for village, household, and individual to account for clustering and repeated measures; penalised splines modelled potential non-linear effects of age. The second tested associations with prior clinical malaria (T1–T2) and carriage at the previous survey visit (T1). The third, also at T2, focused on behavioural factors at T2, the period of highest prevalence, to maximise power for detecting risk factors. The second and third models included random intercepts for village and household only.

## Results

### Participants inclusion and follow-up

670 participants enrolled: 314 (47%) from 45 households in Kati; 356 (53%) from 53 households in Dire (Fig.S1). In Kati, 63 participants (20%) and 5 households (11%) were lost to follow-up; 251 (80%) completed four visits. In Dire, 66 participants (18.5%) and 1 household (2%) were lost, 28 new participants joined during T1 and 243 (68%) completed all four visits.

### Participants description

The mean household size was approximately five participants in both sites. 53.8% of participants were under 15 years in Kati and 48% in Dire. 50.6% were females in Kati and 55.2% in Dire (Table 1).

**Table 1:**
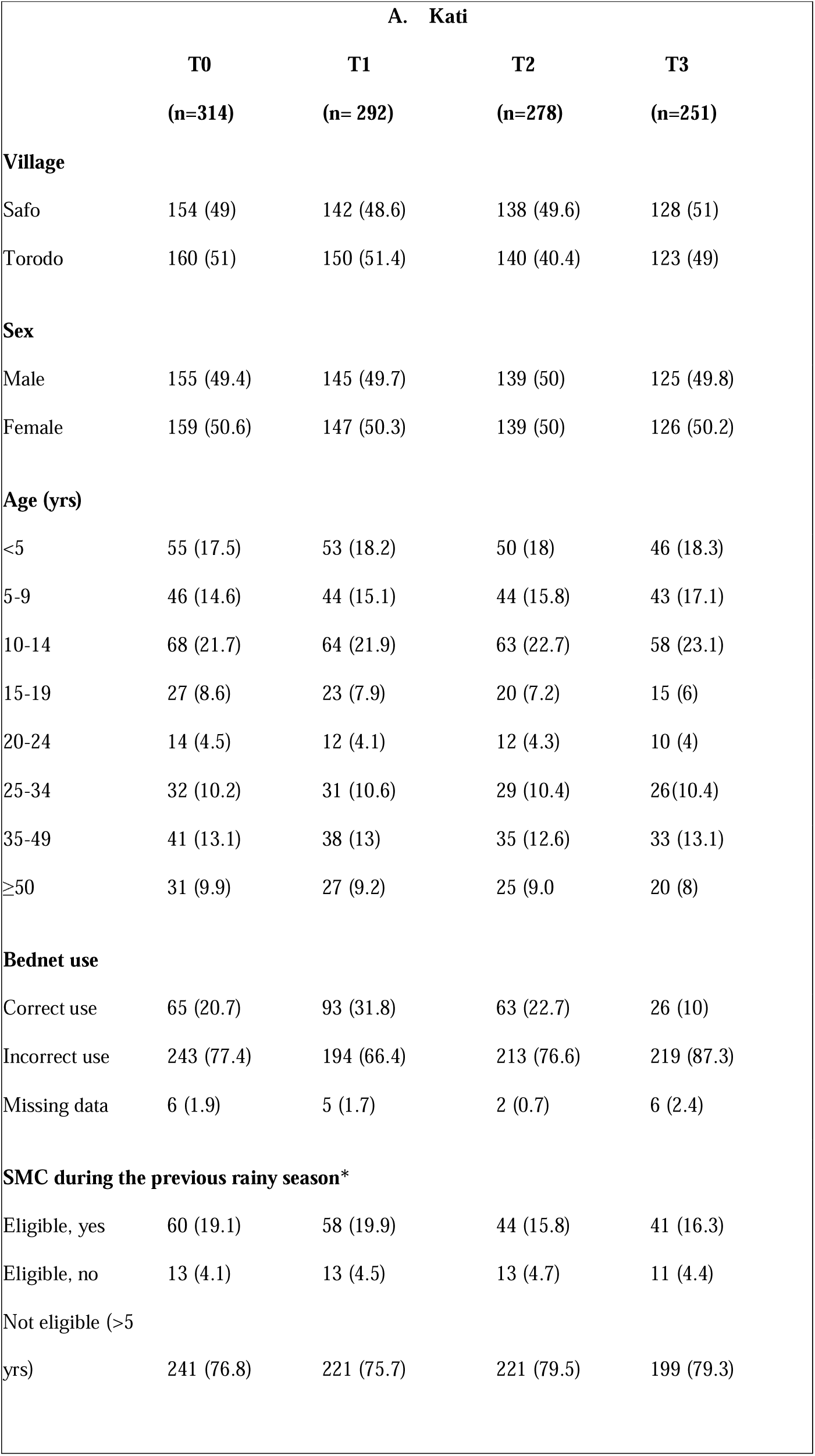
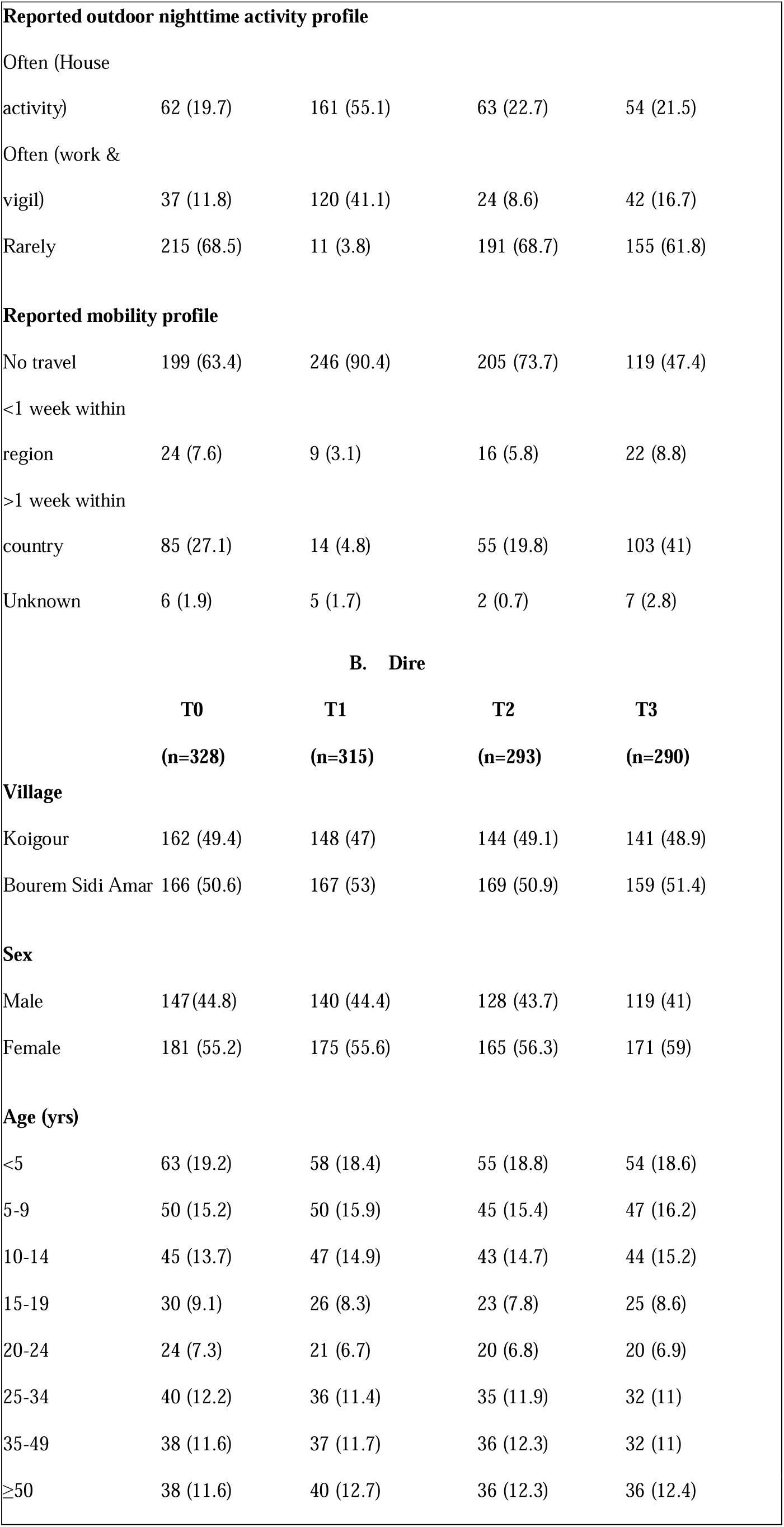
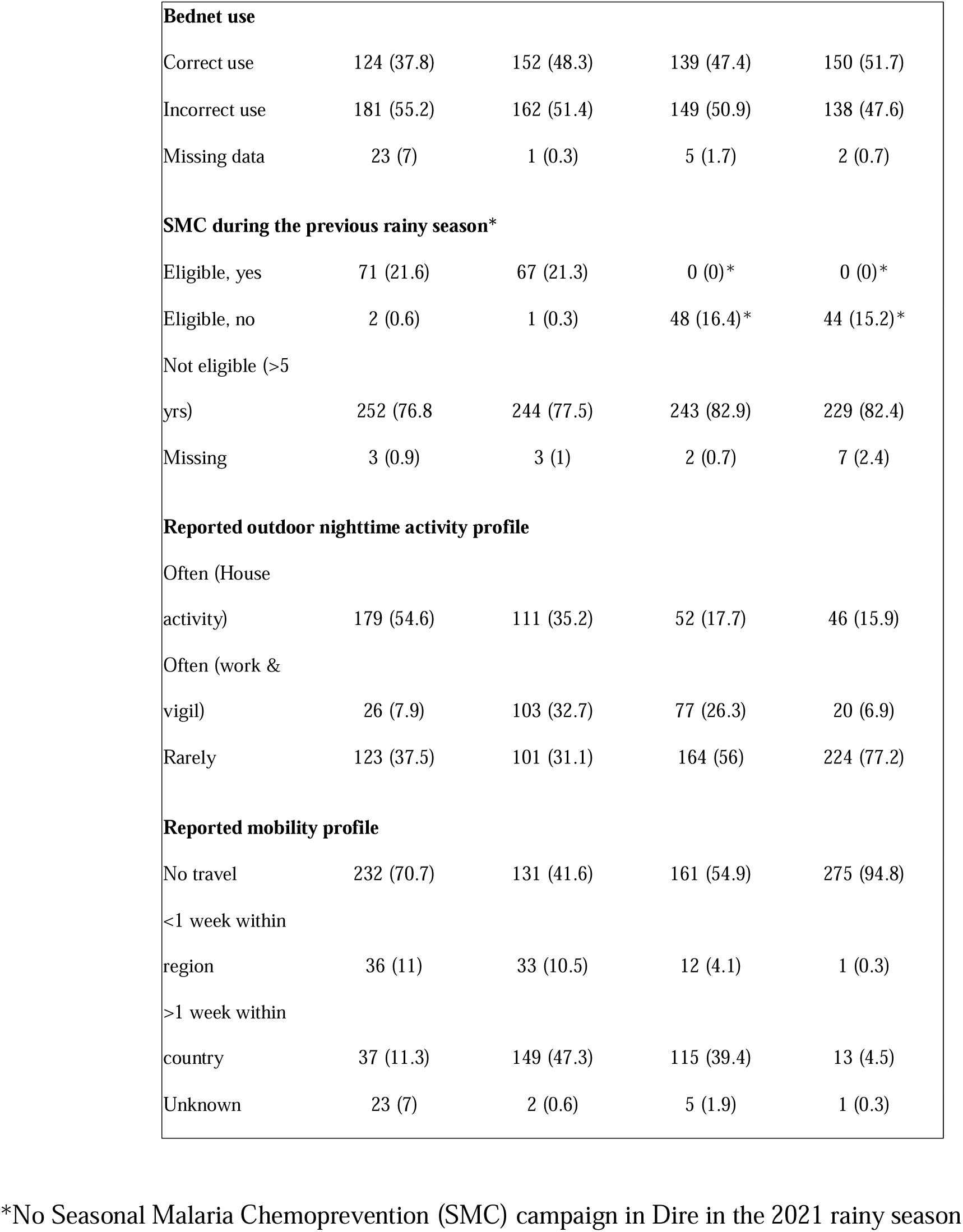
Socio-demographic characteristics and Health behaviour of cohort participants linked to malaria. N (%)

At baseline survey (dry season 2021), reported SMC participation among eligible children (<5 years) was 82% in Kati and 97% in Dire for the 2020 SMC campaign (Table 1). Post SMC (2021, T2), reported participation was 77% (44/57) in Kati. SMC was not implemented in Dire (Table 1). Reported correct bed net use ranged from 10% to 31.8% in Kati and 37.8% to 51.7% in Dire (Table 1). Household nighttime activities were more common than work/vigil, except at T2 in Dire (Fig S2). Most reported no travel (41.6%–94.8%); among those who did, trips lasting more than one week were more common (Table 1, Fig. S3).

### Clinical malaria incidence

During follow-up, 89 clinical malaria cases were detected passively among cohort participants in Kati (Table S1). During the wet season 2021 (June-October, 894mm of rainfall) incidence began to rise in July. Incidence declined with SMC rounds then rebounded in subsequent weeks (Fig. 1). Incidence was highest among ages 20–24 years (Fig. S4). The analysis of total confirmed cases from the village recorded at the local health facility showed a different pattern, with highest incidence among children aged 5-9 years and decreasing with increasing age (Fig. S4).

**Fig. 1.**
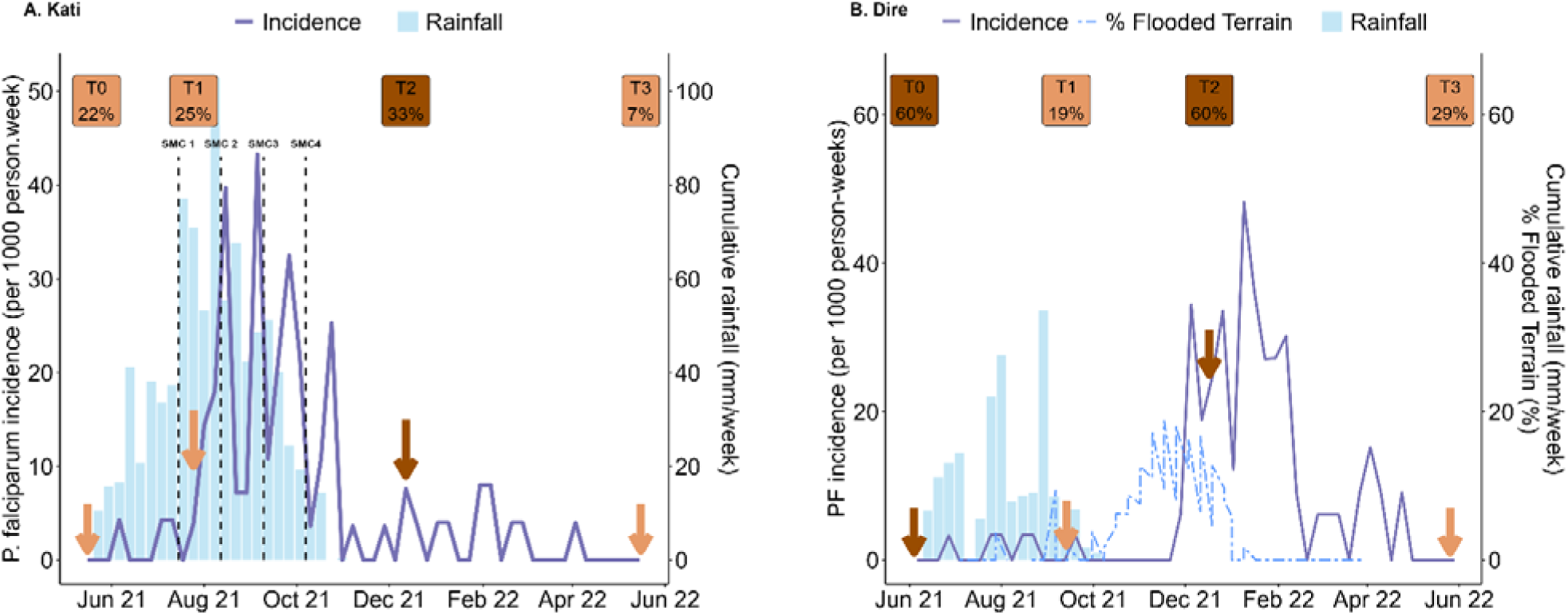
Weekly clinical malaria incidence, rainfall, and flooding during cohort follow-up in Kati (A) and Dire (B). Vertical dashed lines indicate the timing of the four Seasonal Malaria Chemoprevention (SMC) rounds in Kati: SMC1 (15 July 2021), SMC2 (12 August 2021), SMC3 (9 September 2021), and SMC4 (7 October 2021). No SMC was implemented in Dire in 2021. Arrows indicate the timing of malaria prevalence surveys, and shaded boxes highlight survey periods and corresponding prevalence (%).

In Dire, 127 clinical malaria cases were detected passively among cohort participants (Table S1). Incidence rose more intensely during the Niger river flooding period (December-march, Fig. 1) than during the wet season (July-October, 181mm rainfall only). Ages 5–24 years had the highest incidence during the peak period (T2–T3, Fig. S4). The T2 survey coincided with the start of the high-transmission period in one Dire village but not the other (Fig. S5).

### P. falciparum prevalence

In Kati, qPCR prevalence was highest (33.3%) at the end of the wet season (T2, Fig. 1A, Fig. S6). Ages 10–24 years old had the highest prevalence (>40%) during the wet season (T2, Fig. 2A). During dry low-transmission (T0, T3) highest prevalence was among 15-19 year olds (Fig. 2A). In Dire, prevalence varied strongly from one survey to the next. From 60% at T0, it dropped to 19% at T1, to 60% at T2 and 29% at T3 (Fig 1B, S6). Children <5 years had the lowest prevalence at T0 and T1, but without SMC levels were similar to other age groups at T2 and T3 (Fig. 2B). RDT Sensitivity compared with qPCR ranged from 6–28% in Kati and 21–32% in Dire (Table S2). Subpatent infections (i.e. qPCR-positive, RDT-negative) were more frequent in participants older than 20 years (Fig. 2B)

**Fig. 2.**
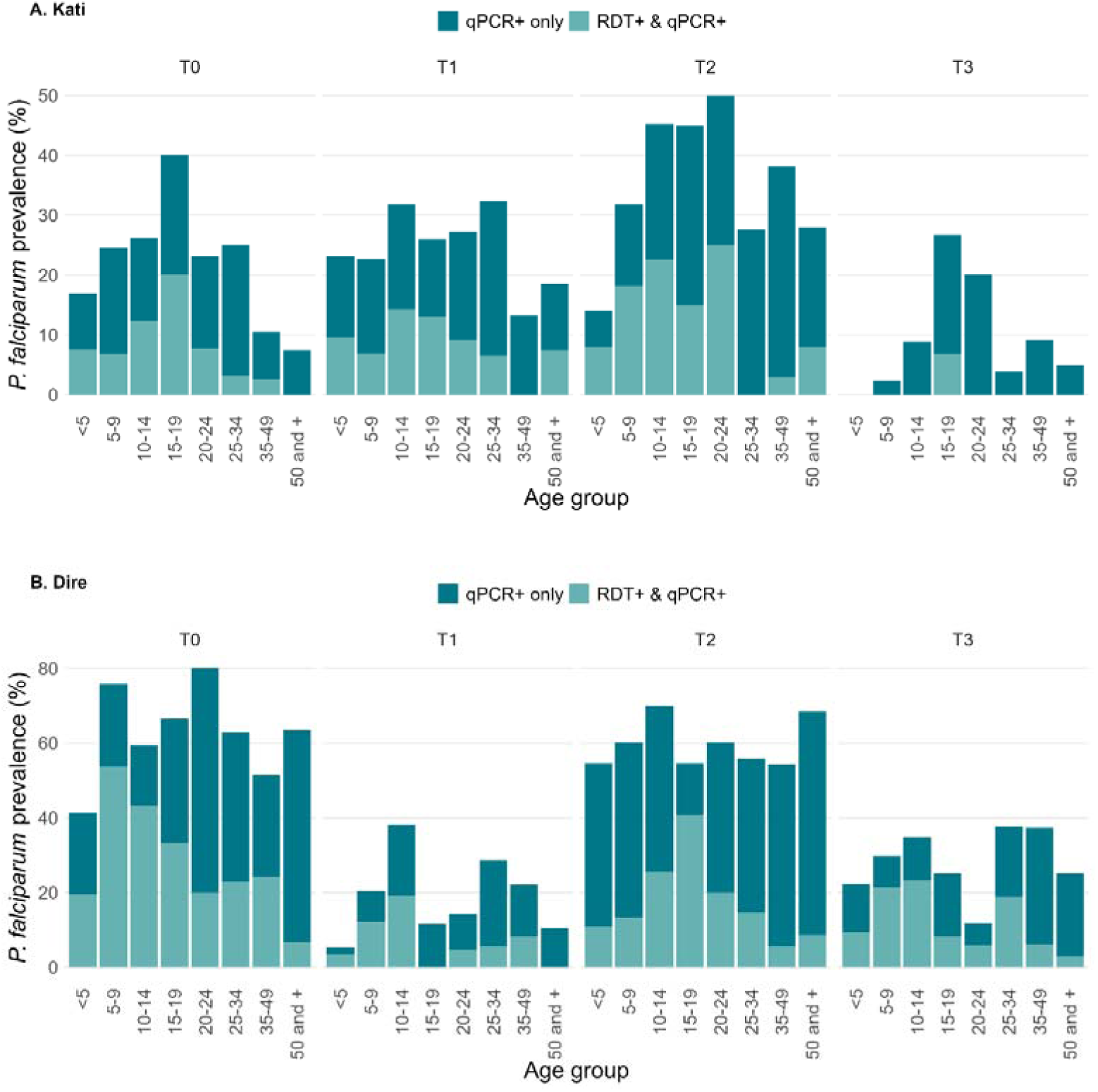
*P. falciparum* prevalence by age group among participants in Kati (A) and Dire (B)

### *P. falciparum* parasitaemia

Over 50% of infections had parasite densities below 100 parasites/μL; at T3 in Kati, all infections were below this (Fig 3). In Kati, median densities remained below 100Lp/μL across all age groups, with slightly higher median in the 10–19 age group at T2 (Fig. 4) but not statistically significant at any visit (Kruskal–Wallis test, *p* > 0.05) (Table S3). In Dire, median densities exceeded 100Lp/μL at T0 (5–14 year olds), T2 (15–19 year olds) and T3 (5-9 year olds) (Fig. 4), statistically significant at T0 (*p* = 0.0006) and T3 (*p* = 0.0391) (Table S3).

**Fig. 3.**
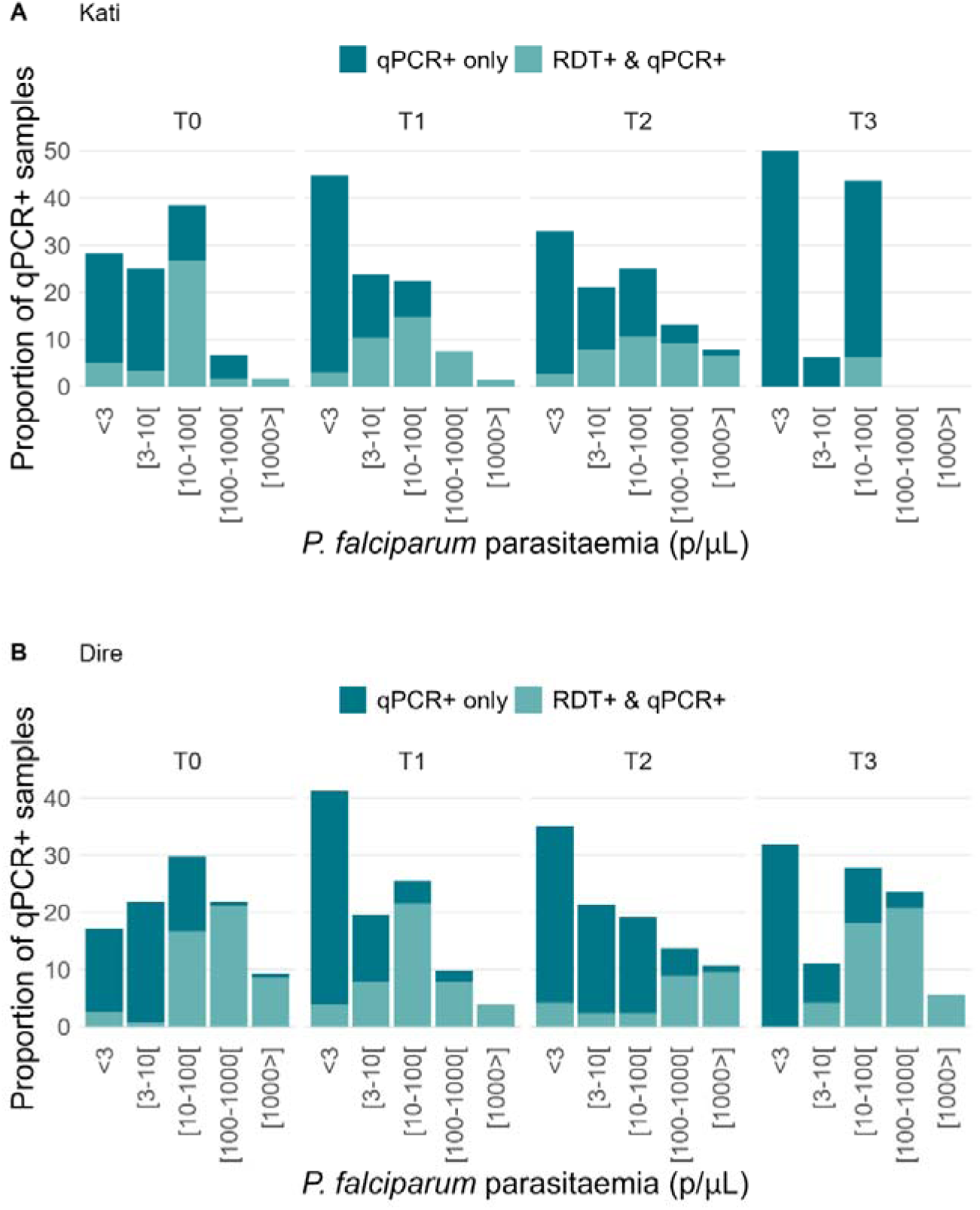
Distribution of *P. falciparum* parasitemia (p/µL) in qPCR-positive samples during surveys in Kati (A) and Dire (B).

**Fig. 4.**
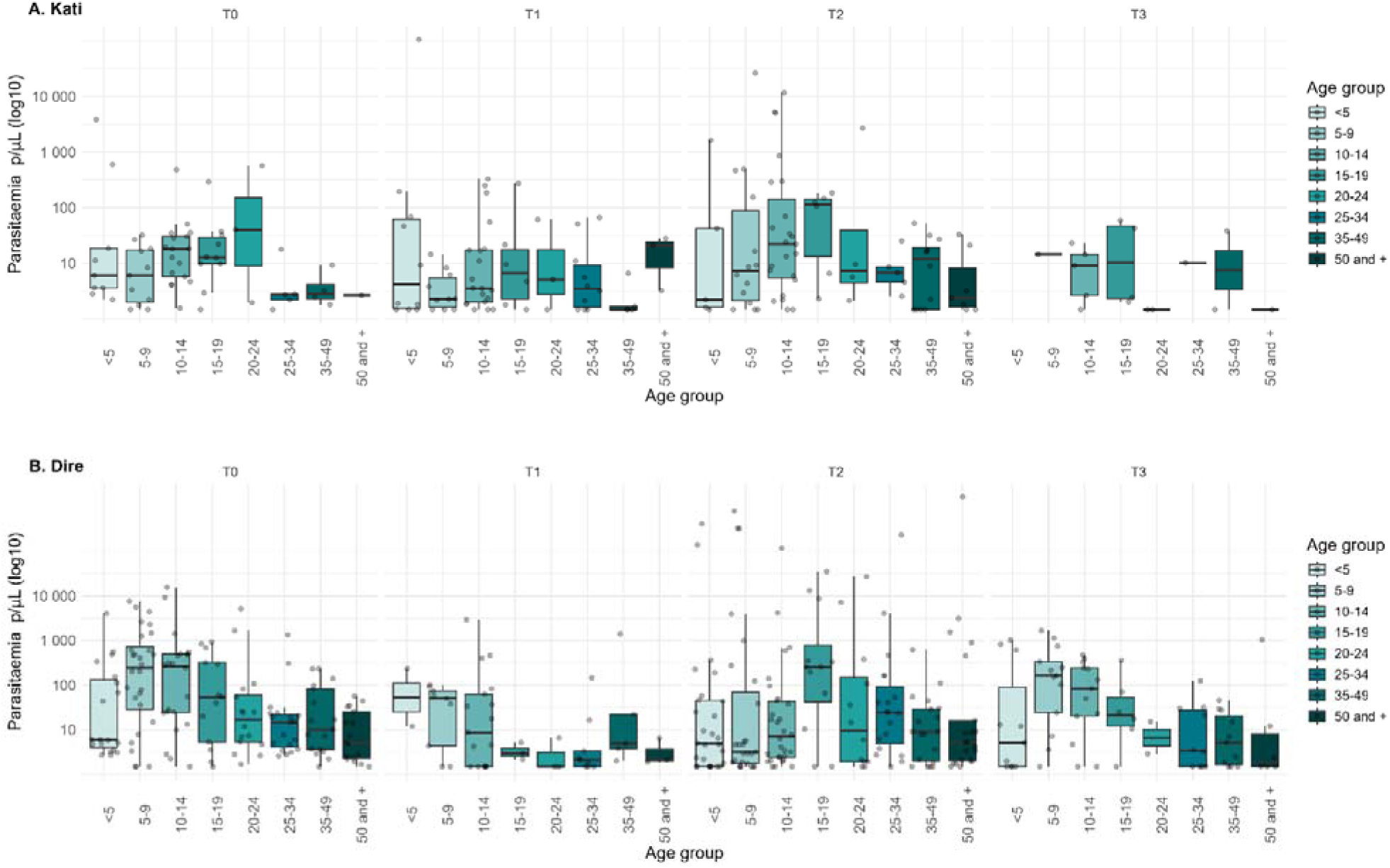
*P. falciparum* parasitemia by age-groups in qPCR-positive samples during surveys in Kati (A) and Dire (B).

### Multiplicity of infection

We obtained 300 genetic barcodes from qPCR positive survey participants of Kati (n=92) and of Dire (n=208). The proportion of polyclonal infections was slightly higher in Kati (71.7%) than in Dire (66.8%). Median eMOI was higher in Kati (1.37) than in Dire (1.25) (Fig. 5), indicating a predominance of polyclonal infections in both sites, with slightly more complex infections in Kati. The difference across age groups was not statistically significant in both sites (Kruskal–Wallis test, *p* > 0.05). Median eMOI values remained above the polyclonal threshold for all age groups, indicating a predominance of polyclonal infections, except Kati 20-24 year group (Fig. 5). Children under 5 years had eMOI similar to older age groups. In Kati, the median eMOI was highest in 10–19-year-olds, while in Dire, eMOI peaked in the 20–24-year-olds. Overall, these patterns indicate more complex infections in under-5s and 10-24 year olds.

**Fig. 5.**
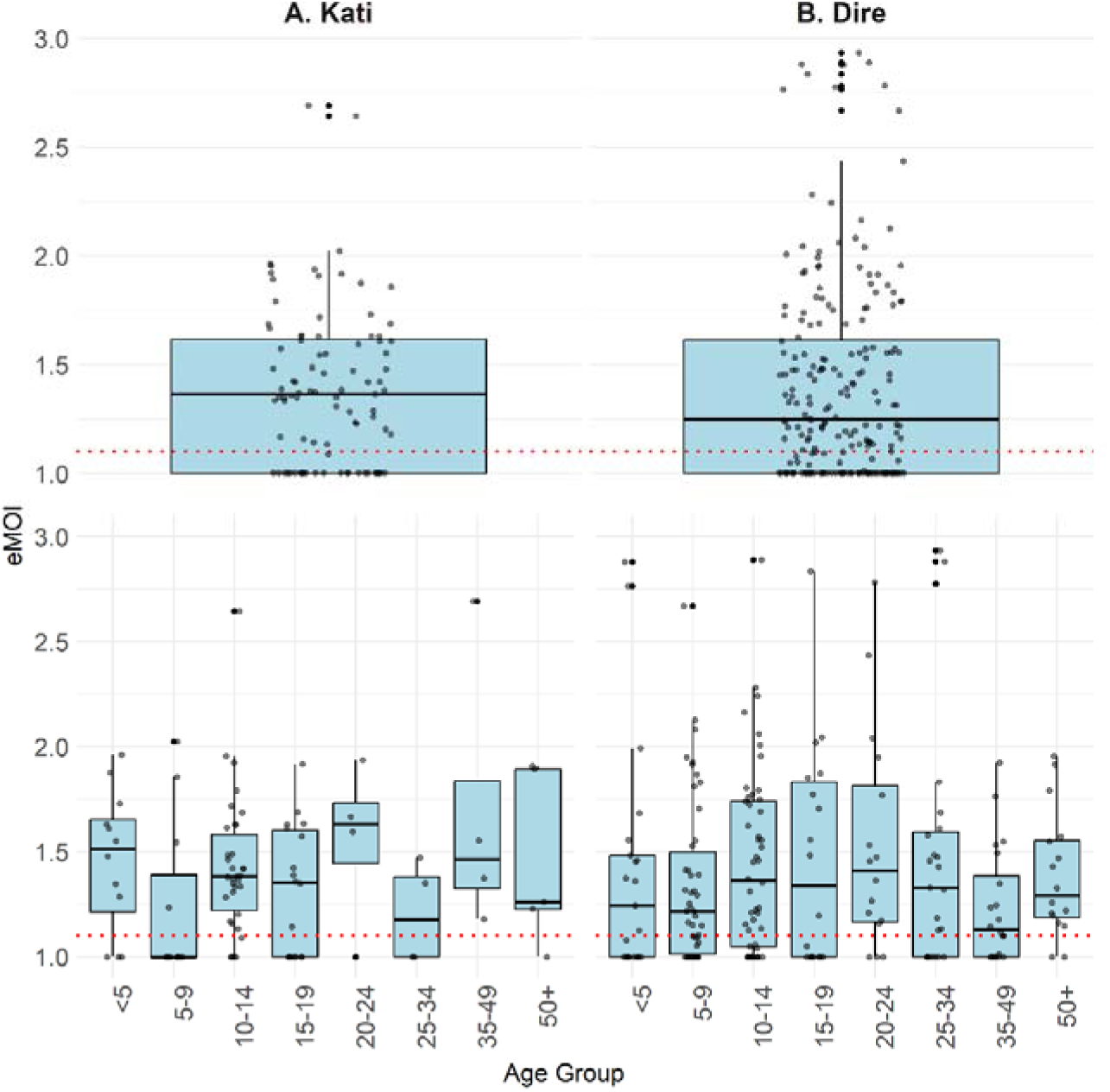
eMOI distribution overall and by age among qPCR-positive participants in Kati (A) and Dire (B). The red dotted line indicates eMOI = 1.1, threshold used to classify polyclonal infections.

### Factors associated with *P. falciparum* carriage

In Kati, odds of infection increased from dry season (T0) to end of wet season (T2, OR = 2.43, 95% CI: 1.49–3.96), then dropped by T3 (OR = 0.13, 95% CI: 0.06–0.29, Table 2). In Dire T0 and T2 were similar, and T1 (OR = 0.10) and at T3 (OR = 0.23, Table 2). There was no sex association at either site. Age- patterns varied by site and survey. In Kati, age effects were most evident at T2 and T3, where children under 5 years had lower odds of infection (eg OR = 0.57, 95% CI: 0.35–0.94 T2), with risk increasing steadily with age (Additional File1: Fig. S7). In Dire, non-linear associations were observed at T0 and T1, with significantly lower odds among children under 5 years (eg OR = 0.70, 95% CI: 0.50–0.97 at age 5 in T0), and a peak in 10–19 years olds (e.g., OR = 1.64, 95% CI: 1.07–2.50 at age 19 in T0) (Additional File1: Fig. S8).

**Table 2:**
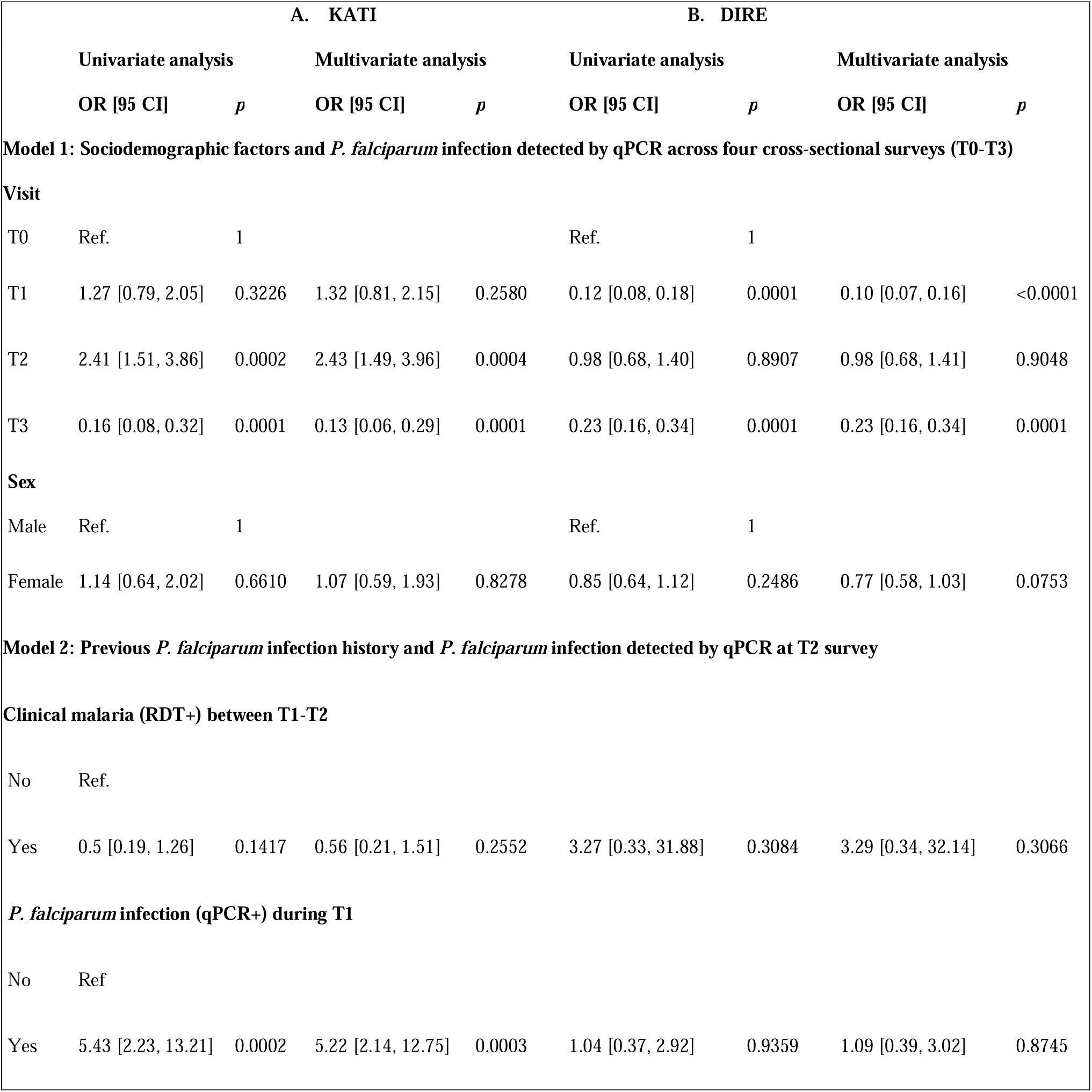
Odds Ratios (OR) estimates and 95% Confidence Intervals (CI) based on GAMM.

At T2 (Kati), non-SMC eligible participants had much higher odds of infection than eligible children who received SMC (OR 16.29, 95% CI 3.32–79.79, p = 0.0006; Additional File1: Table S4). In Dire, travelling more than 1 week within the country lowered odds of infection than non-travellers (OR 0.38, 95% CI 0.18–0.80, p = 0.010). No link to bed net use or outdoor night activity at either site (Additional File1: Table S4)

Clinical malaria (RDT between T1- T2) was not associated with infection at T2 (Table 2). Prior qPCR positivity (T1) was strongly linked to infection at T2 (OR = 5.22, 95% CI: 2.14–12.75, *p* = 0.0003) in Kati, but not in Dire

## Discussion

This study provides a comprehensive assessment of *P. falciparum* infections in two different epidemiological settings of Mali. In central Mali, Kati followed “classic” Sahelian seasonality and received routine SMC for children under 5. The distribution of *P. falciparum* infections in Kati presented significant changes compared to pre-SMC era. Under-5s appeared well protected, except at T1 which took place at the same time as SMC round 1 and showed a higher prevalence, suggesting new infections since the onset of the rainy season. Age groups older than 5 years displayed the expected pattern of increasing prevalence (from 5 to 24) and increasing parasite densities (from 10 to 19), as well as higher eMOI. In the general village population, clinical malaria incidence was highest in children aged 5-9 years, immediately above SMC-eligibility.

In Northern Mali, malaria seasonality of 2021 in Dire reflected a stronger influence of Niger River flooding than of rainfall. The distribution of *P. falciparum* infections followed a complex pattern. The baseline survey did not take place in the midst of the low transmission season but shortly after the end of the major flood-related malaria peak, which explains the high prevalence. RDT-positive infections represented 30% and received treatment. The following low transmission period from June to September might explain the following low prevalence at T1 in September. T2 corresponded to the start of the flood-related malaria peak, accompanied with high prevalence again. Clinical malaria incidence appeared northern Mali, presenting a complex pattern of high prevalence, are widespread in Mali and persist across age groups and seasons. Three main groups emerged: under-5s well protected with SMC but quickly exposed otherwise; children and youth (5–24 years), with the highest prevalence and parasite densities; and adults over 25, frequently infected at lower densities. Most Infections are undetected by RDTs and less likely to be treated, as they frequently cause little to no symptoms thus contributing to the transmission reservoir. Genetic analysis showed polyclonal infections were highest in adolescents and young adults, but still elevated in under-fives (SMC eligible), indicating that age groups beyond current SMC coverage also carry complex infections and contribute substantially to the parasite reservoir.

Transmission dynamics differed sharply between sites. Kati follows a typical rainfall-driven Sahelian profile, with low-density infections after rains. This is consistent with earlier analyses showing strong spatio-temporal variability of incidence linked to rainfall, river flow, and land use, with hotspots persisting even during the dry season[16]. The low clinical incidence suggests strong immunity. Dire patterns were dominated by rainfall and Niger River flooding, plus irrigated agriculture causing a January-March high transmission season [17,28].

Although Kati receives more rainfall, prevalence was consistently higher in Dire. Combined with higher parasite densities and lower eMOI compared to Kati, this suggests lower population immunity. This is consistent with the intensification of malaria transmission after the development of irrigated agriculture, and a population historically less exposed. The absence of SMC in Dire reinforced infections in children <5 during high transmission (T2). A cohort effect may be visible related to (i) the treatment of RDT-positive cases during surveys and (ii) improved access to treatment of clinical malaria for cohort participants, which may have contributed to lower prevalence in T1 and T3 surveys. These observations complement previous studies showing Dire’s bimodal transmission patterns driven by rainfall and flooding[17], with our cohort adding that during the flood-driven peak, asymptomatic infections are widespread across all ages, with reinfections dominating in the absence of SMC.

The high polyclonal infections imply ongoing exposure to diverse parasite strains, even among asymptomatic carriers, explaining sustained transmission despite lower clinical incidence. Median eMOI of 1.37 in Kati and 1.25 in Dire indicates most infections are biclonal, with one dominant and one minor clone. These intermediate values suggest cotransmission rather than is the main source of polyclonality consistent with limited transmission diversity in these settings.

Population analyses in Kati show that SMC reduces community-level incidence [29]. Our cohort confirms protection for under-5s. Our results also show that those aged 10 to 24 had the highest wet season prevalence (T2), but also the highest parasite densities and multiplicity of infection. These results complement findings from Senegal, where SMC extends to age 10 years, SMC-eligible children have the lowest prevalence, while adolescents and young adults aged 15-24 years carried the greatest burden[7]. Our results suggest that those who receive SMC from infancy become key contributors to the reservoir after 5 years with high prevalence but also as individually good hosts (highest densities and eMOI). In Senegal, children immediately older than the SMC eligibility threshold presented the highest clinical malaria incidence not seen here. Mali with SMC stopping at age 5, youths and young adults already accounted for a large share of infections despite earlier exposure. Compared with Senegal, Mali showed both higher overall prevalence reflecting more intense transmission and persistent barriers to timely access to care outside study conditions.

Our findings have important public health implications. SMC is highly effective in under-fives but leaves a large untreated reservoir in older children and adults. The Dire experience highlights the rapid reversal when SMC is interrupted[30]. Beyond diagnostics, baseline MOI and genetic diversity provide essential metrics which could be instrumental in completing the assessment of interventions such as SMC, as they capture shifts in transmission intensity and parasite population structure[31]. In addition, expanding SMC eligibility, exploring school-based delivery, and evaluating focal mass drug administration are potential strategies to address transmission beyond current control strategies. Strengths of this study include the use of sensitive molecular diagnostics, high-resolution longitudinal follow-up, and rare population-based data from both Central and Northern Mali, where security challenges have often limited research. Limitations include modest sample sizes in some strata, a cohort effect from repeated survey contacts, and enhanced access to free diagnosis and treatment for participants, which may have reduced chronic carriage compared with non-participants. Differences in cohort structure and survey-based treatment of RDT-positives between sites also shaped infection dynamics, but these contrasts created a valuable natural experiment to assess the combined effects of ecology and intervention coverage.

## Conclusion

*P. falciparum* infections are highly prevalent in the general population of two regions of Mali. They concentrate on children from 5 years older than SMC eligibility and youth up to 24 years, but are also present in adults. The historical paradigm of the highest incidence in the youngest children is altered by SMC, and these changes should be taken into account to design additional strategies. Indeed, while vaccines are rolled out to protect the youngest children, those older are supporting persistent *Plasmodium* transmission. Strategies targeting asymptomatic carriers are likely required to further decrease Plasmodium transmission and continue progress towards elimination in seasonal settings.

## Data Availability

All data produced in the present study are available upon reasonable request to the authors

## Supplementary information

**Additional file includes** Figs. S1- S8 and Tables S1-S4

## Declarations

### Ethics approval and consent to participate

The study protocol was approved by the Ethics Committee of the University of Sciences, Techniques and Technologies of Bamako, Mali (protocol No. 2020/297/CE/FMOS/ FAPH).

### Consent for publication

Not applicable

### Availability of data and materials

The datasets used and/or analysed during the current study are available from the corresponding author on reasonable request.

### Authors’ contributions

JL, IS, JG, and E-HKCB designed the study, with contributions from SR, MCi, MKBD and AD. JL, IS, E-HKCB, JG, MKBD, MCi, AK, EL, AC, and CL’O designed the methodological approach, study procedures, and tools. AD, ODAP, MC, PK, BKam, MBK, PK, AZ collected data and samples. CL’O, MM designed and conducted the PCR assays. JL and BKa designed the statistical analysis plan and assessed and verified the data. BK conducted the analysis excepted for the eMOI analysis conducted by EBB. JL, BKa and EBB interpreted the findings. BKa and EBB created the figures. BKa wrote the first draft of the manuscript. All authors reviewed and revised the manuscript and approved this version of the Article for publication.

## Acknowledgements

We sincerely thank all participants, community leaders, and the residents of Safo, Torodo, Koigour, and Bourem Sidi Amar for their engagement and active role in advancing malaria control efforts. Their generosity and cooperation made this work possible. We extend special appreciation to the community guides, and health post staff for their dedication, and to the research assistants whose commitment was essential to the success of this study.

