## Supplementary material for "*Plasmodium falciparum* Carriage in central and northern Mali amid Seasonal Malaria Chemoprevention implementation": Fig. S

**ADDITIONAL FILE 1**

**Methods S1: Generating profiles from questionnaire data using PCA and HCPC**

**Mobility and night-time outdoor activity profiles**

Mobility profiles were derived from questionnaire-based mobility data using multivariate analysis. 17 of the **30 mobility-related questionnaire variables** were retained following selection criteria. Observations with missing data for all mobility variables were excluded. Variables with **≥10% missing values** were removed. Among the remaining variables, only those with **<10% missing data and positive responses >10%** were retained to ensure sufficient variability. Additionally, observations with **more than two missing mobility variables** were excluded to limit the influence of incomplete data on the analysis. After applying these criteria, the dataset was reduced from **2,363 to 2,310 observations**.

Each observation corresponded to a unique **individual–visit combination**, defined using an identifier based on record ID and survey visit , to account for repeated measurements within individuals. Mobility variables were collected as **categorical responses**, coded on a **four-level scale (1–4)** corresponding to “Often”, “Occasionally”, “Rarely”, and “Never”, respectively.

A total of **11 night-time outdoor activity variables** derived from questionnaire data were analysed using the **same PCA/HCPC approach** as that applied to mobility variables. These variables were measured using a **five-level frequency scale (0–4)** corresponding to “Never”, “Less than once per month”, “At least once per month”, “At least once per week”, and “Every night”.

For both mobility and night-time activity datasets, **Principal Component Analysis (PCA)** was performed on completed datasets. Principal components were retained based on **variance explained and interpretability of components**. To identify profiles, **Hierarchical Clustering on Principal Components (HCPC)** was applied to the PCA scores. The number of clusters was set to **three**, based on inspection of the dendrogram and cluster interpretability. All analyses were conducted using **R software**, FactoMineR packages, and **PCAshiny**.

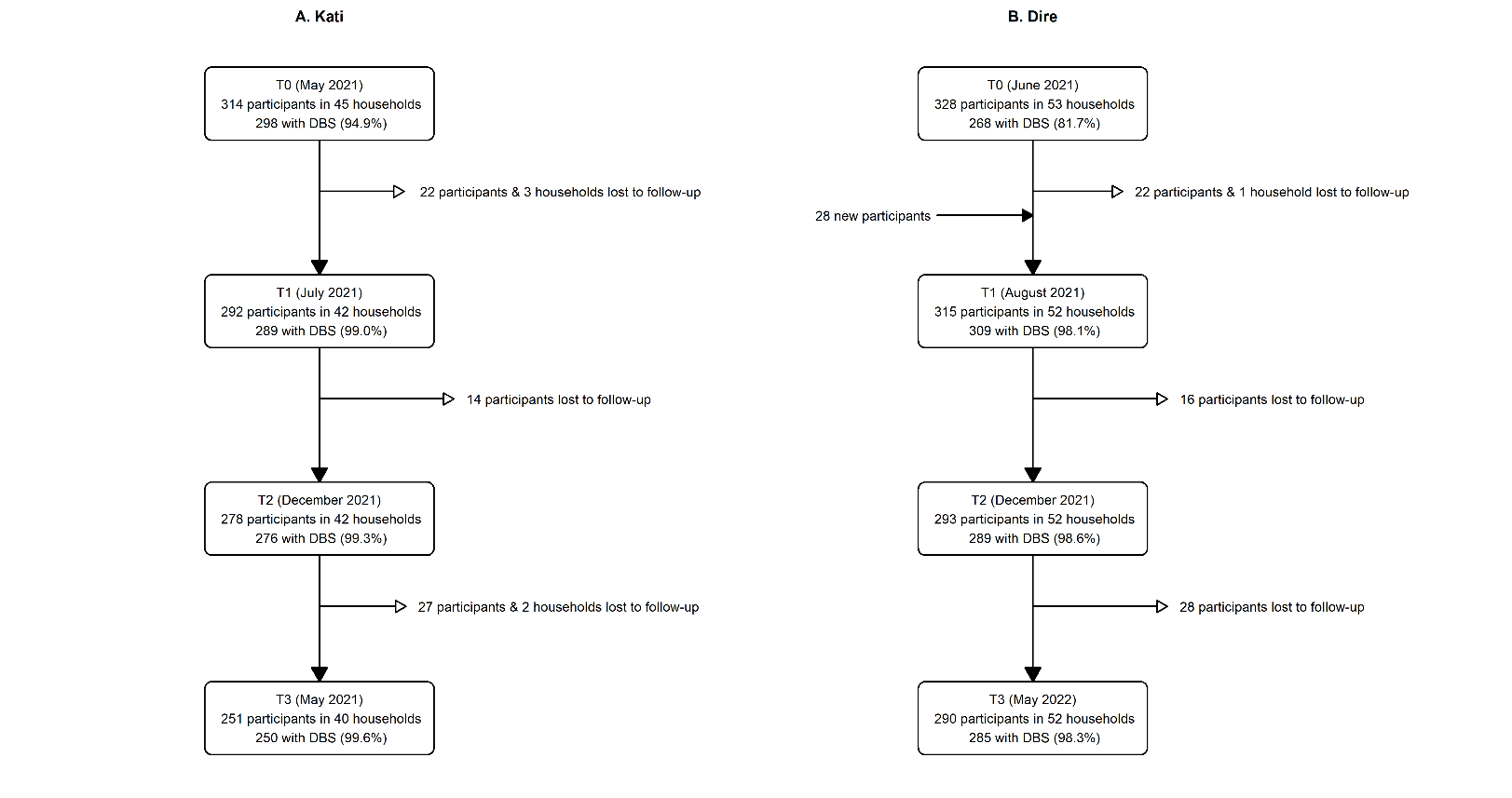

**Figure S1: Flow chart of cohort participants during the follow-up in Kati and Dire, 2021–2022.**

**A.**

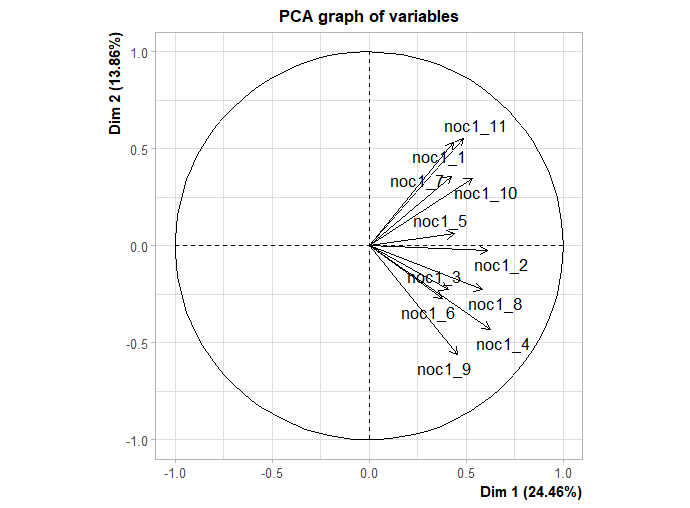

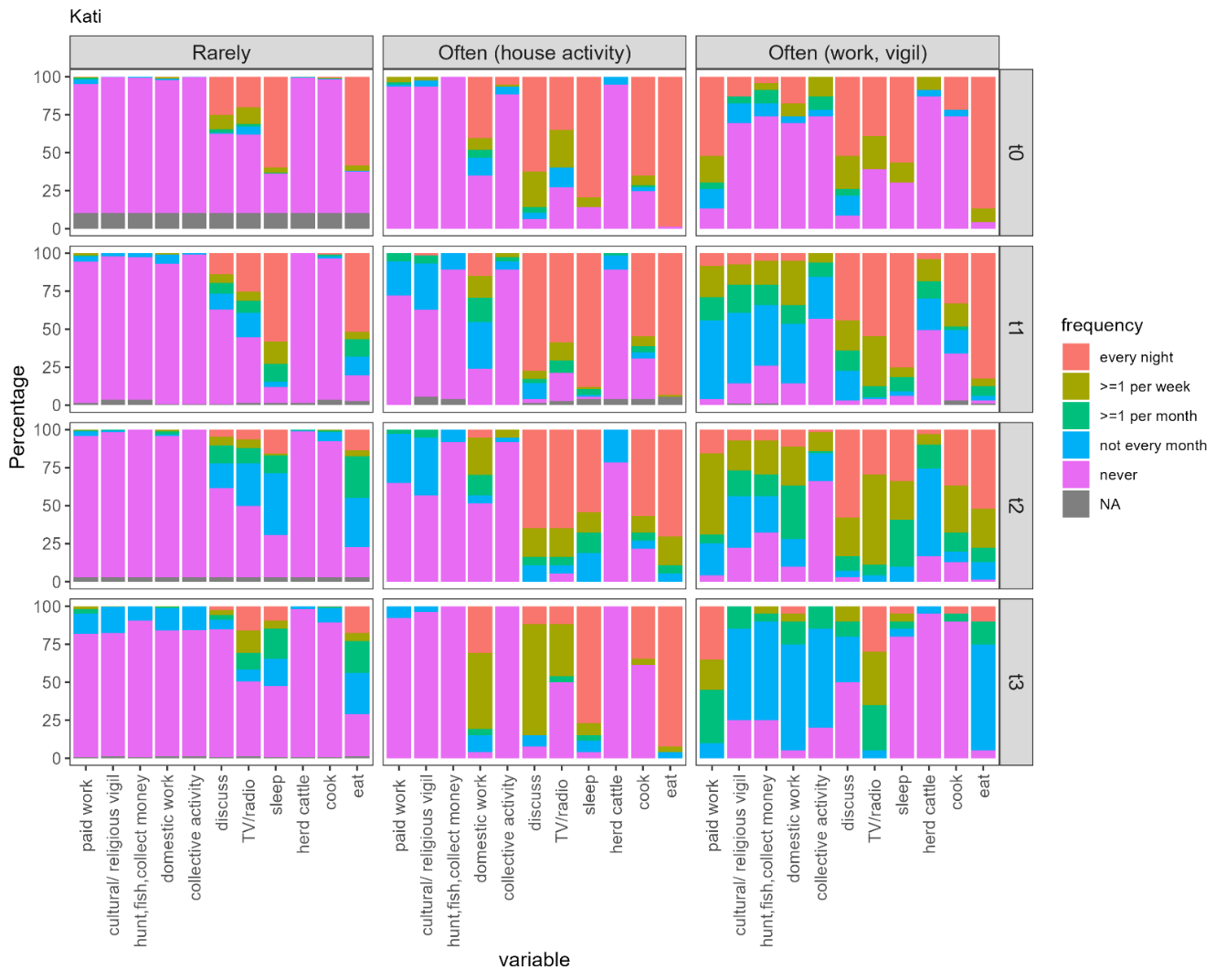

**B.**
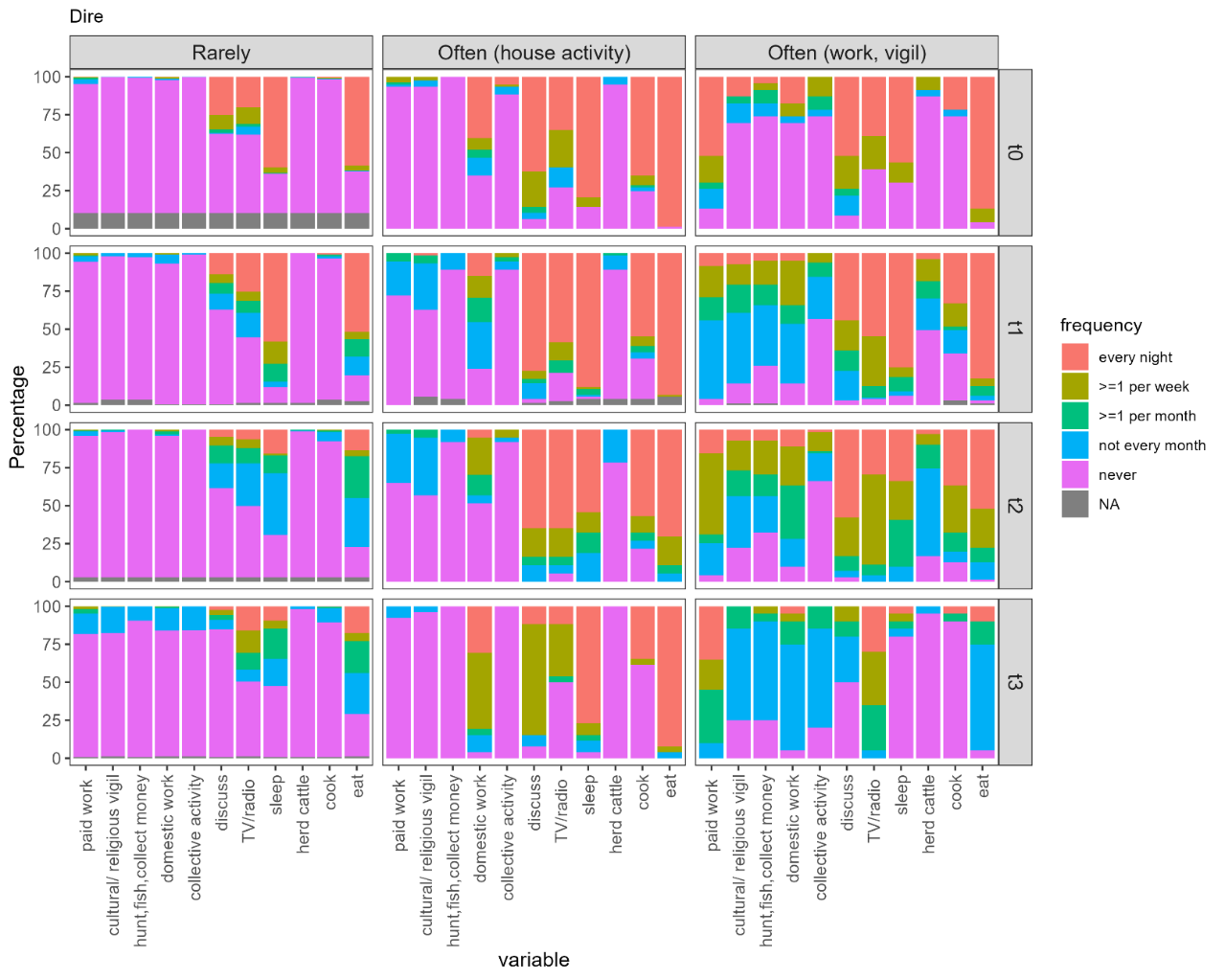

**Figure S2: night-time activity profiles A. correlation between outdoor night-time activity frequency variables (correlation circle) and B. Distribution of response to questions by nighttime activity profile in Kati and Dire**

**A.**

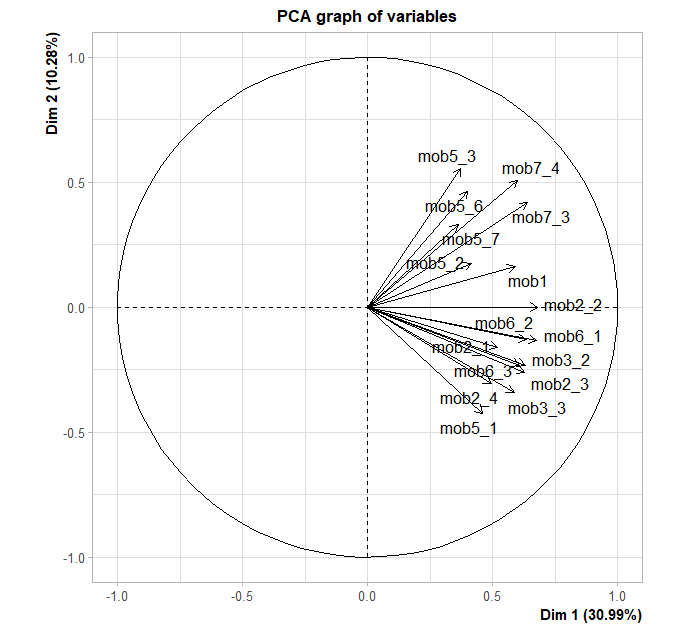

**B.**

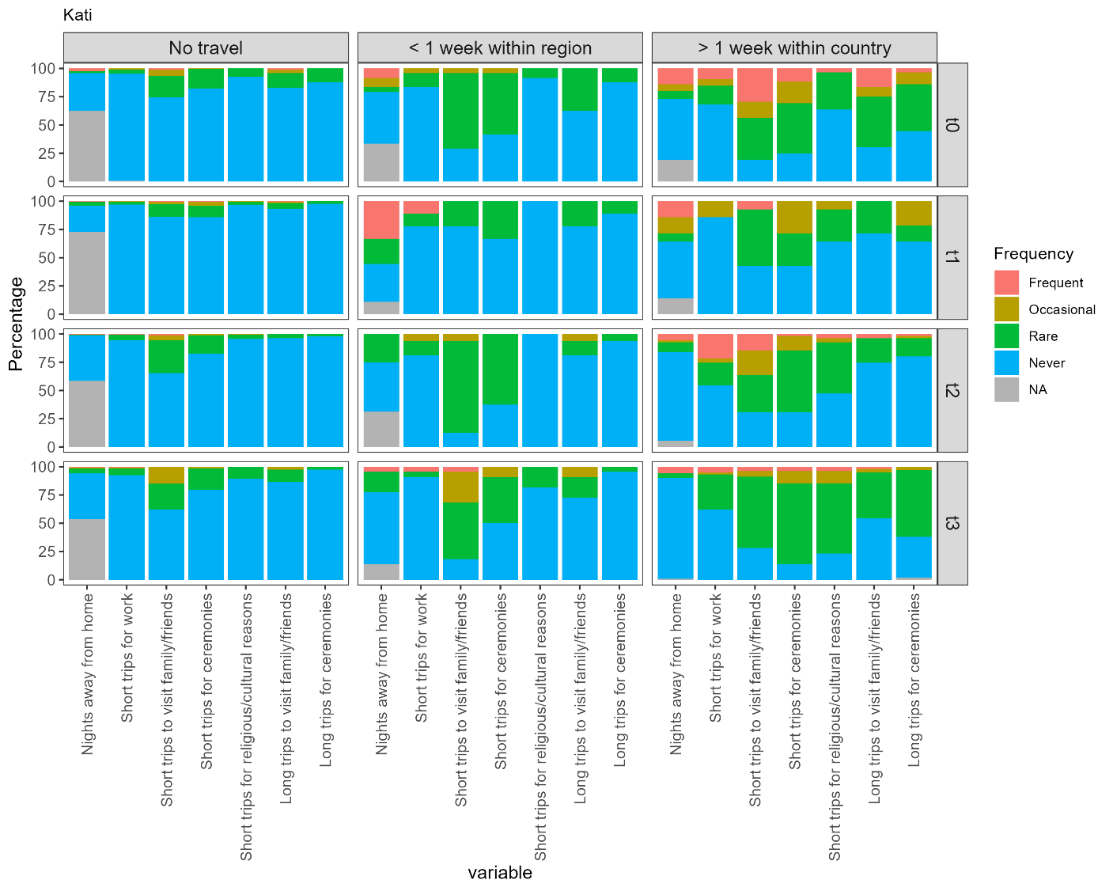

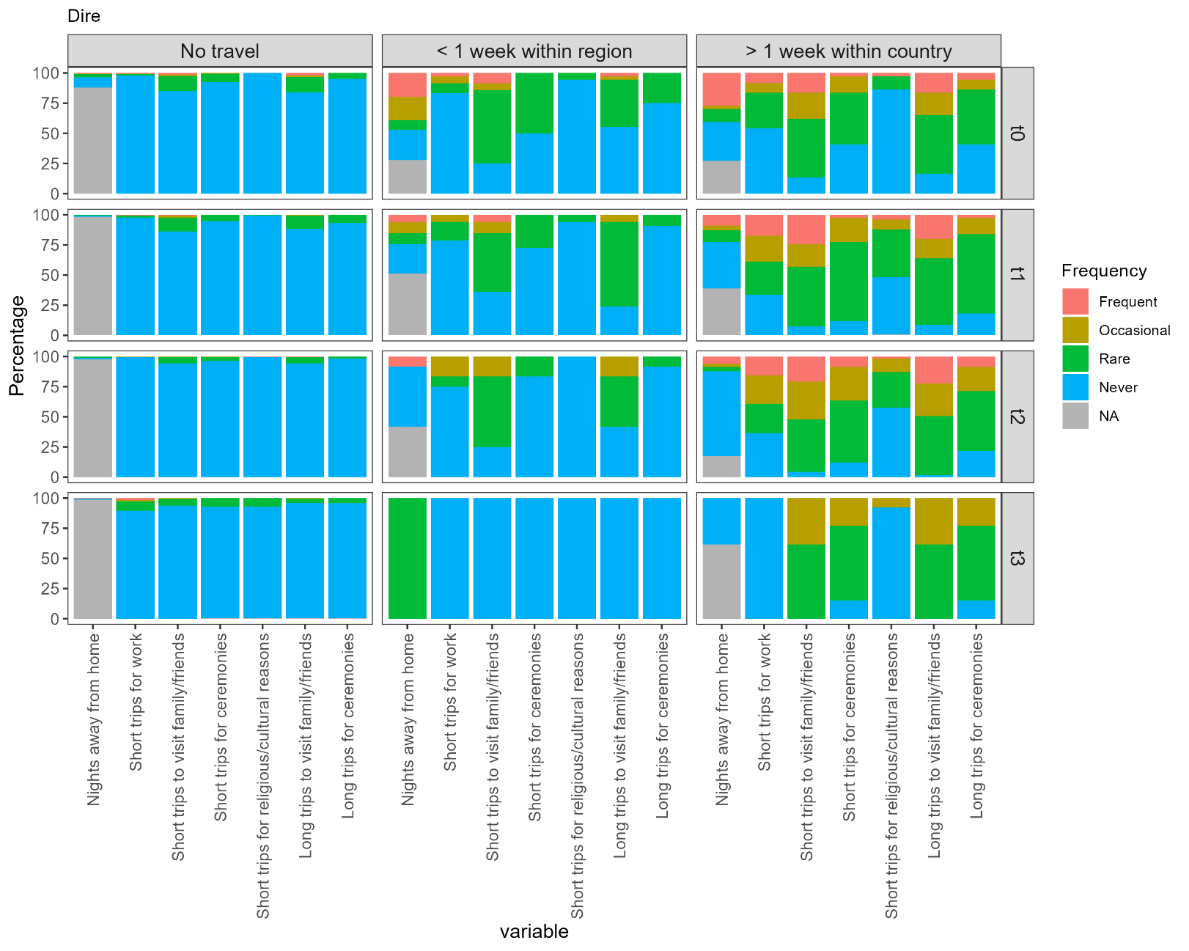

**Figure S3: mobility profiles A. correlation between mobility frequency variables (correlation circle) and B. Distribution of response to questions by mobility profile in Kati and Dire**

**Table S1: Malaria infection of cohort participants. N(%)**

|  | A. KATI | | | | B. DIRE | | | |
| --- | --- | --- | --- | --- | --- | --- | --- | --- |
|  | T0  N=314 | T1  N=292 | T2  N=278 | T3  N=251 | T0  N=328 | T1  N=315 | T2  N=293 | T3  N=290 |
| *P.falciparum* infections (qPCR+) |  |  |  |  |  |  |  |  |
| Yes | 64 (20.4) | 71 (24.3) | 92 (33.1) | 17 (6.8) | 162 (49.4) | 59 (18.7) | 173 (59) | 82 (28.3) |
| No | 234 (74.5) | 218 (74.1) | 184 (66.2) | 233 (92.8) | 106 (32.4) | 250 (79.4) | 116 (39.6) | 203 (70) |
| Missing | 16 (5.1) | 3 (1.0) | 2 (0.7) | 1 (0.4) | 60 (18.3) | 6 (1.9) | 4 (1.4) | 5 (1.7) |
| *P. falciparum* infection (RDT+) |  |  |  |  |  |  |  |  |
| Yes | 26 (8.3) | 26 (8.9) | 46 (16.5) | 1 (0.4) | 85 (25.9) | 31(9.8) | 48 (16.4) | 44 (15.2) |
| No | 277 (88.2) | 263 (90.1) | 230 (82.7) | 246 (98) | 206 (62.8) | 276 (87.6) | 240 (81.9) | 240 (82.8) |
| Missing data | 11(3.5) | 3 (1) | 2 (0.7) | 4 (1.6) | 37 (11.3) | 8 (2.5) | 5 (1.7) | 6 (2.1) |
| *Clinical malaria case recorded since last visit |  |  |  |  |  |  |  |  |
| Yes |  | 4 (1.4) | 72 (25.8) | 12 (4.8) |  | 5 (1.6) | 20 (6.8) | 91(31.4) |
| No |  | 288 (98.4) | 206 (74.2) | 239 (95.2) |  | 310 (98.4) | 273 (93.2) | 199(66.6) |

*Clinical malaria cases recorded since the last visit represent the number of participants present at survey with at least one documented or reported clinical episode between consecutive survey rounds and do not reflect the total number of clinical malaria episodes occurring in the cohort during those intervals. Across all survey periods, Kati recorded 89 cases (4 at T0–T1, 73 at T1–T2, and 12 at T2–T3), whereas

Dire recorded 127 clinical cases (5 at T0–T1, 20 at T1–T2, and 102 at T2–T3).

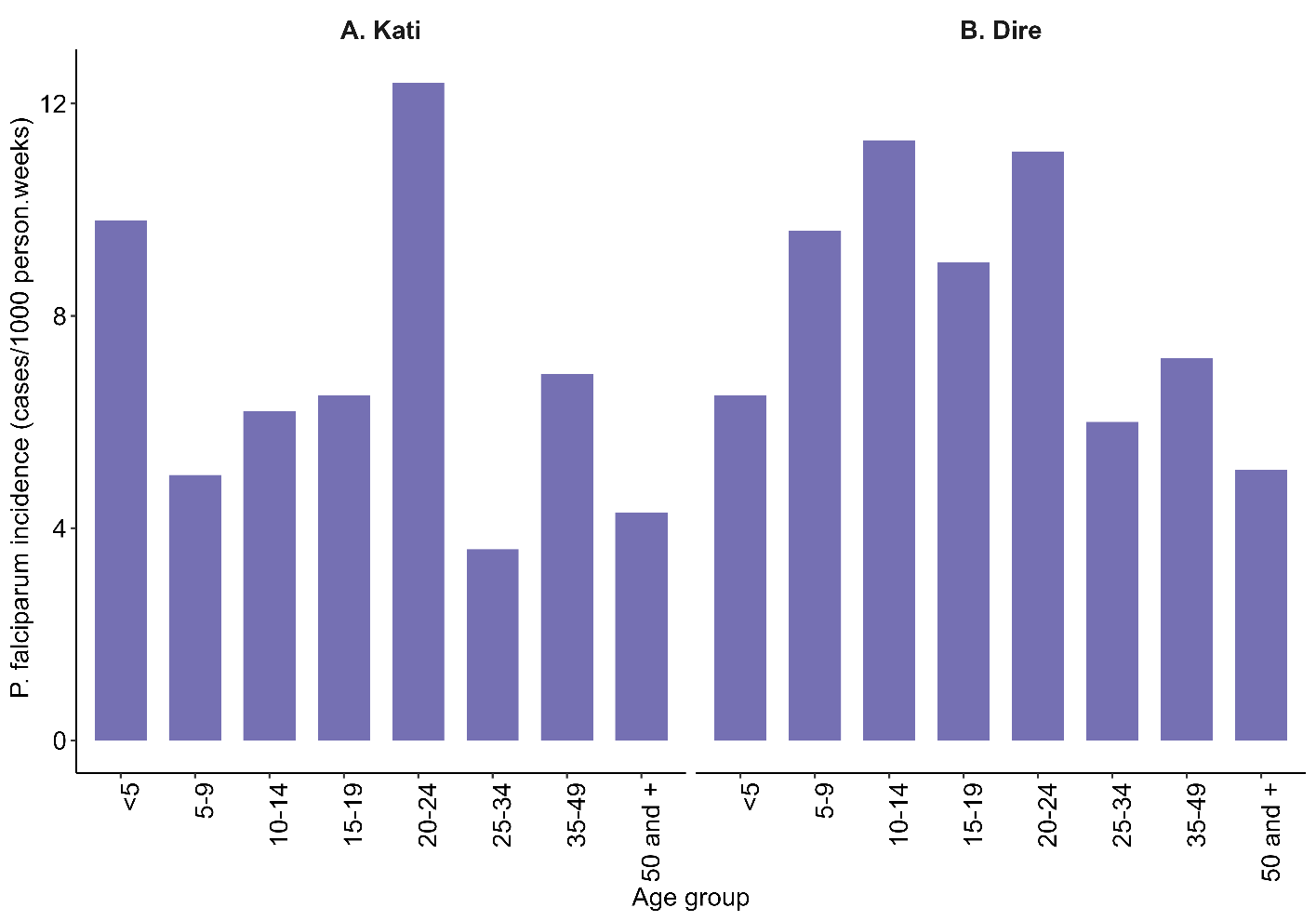

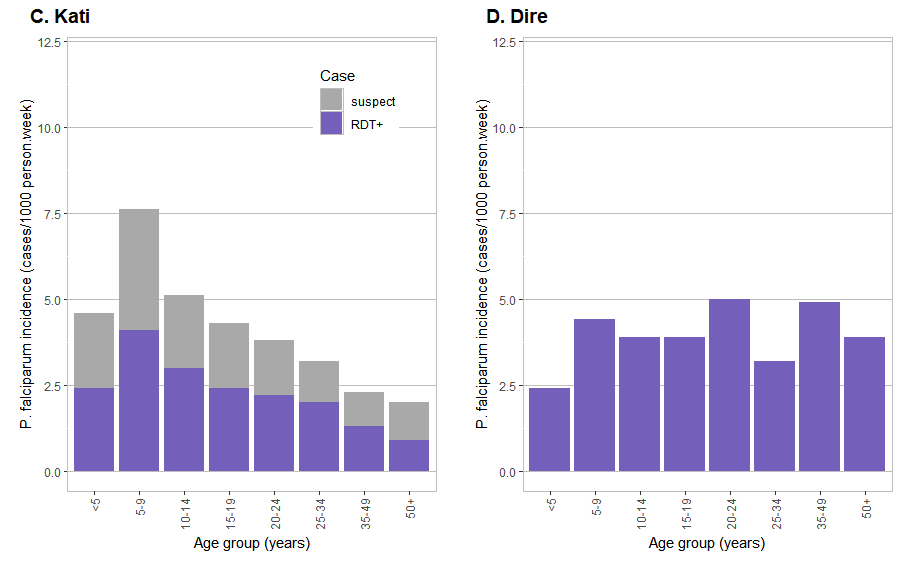

**Figure S4: Age-specific Plasmodium falciparum incidence rates per 1,000 person-weeks over the study period in (A) Kati and (B) Dire among cohort participants (**C) Kati and (D) Dire among the entire village population. Village case counts were obtained from health facility records and village population from project census.

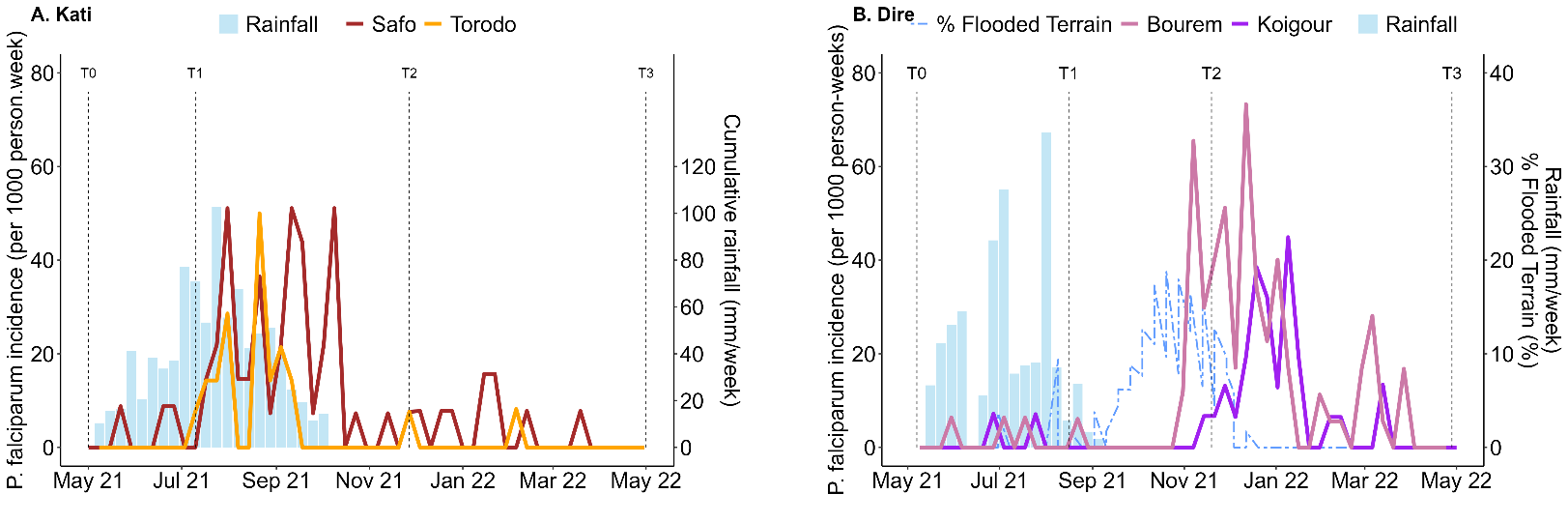

**Figure S5. Weekly malaria incidence and rainfall in (A) Kati villages and ( B)Dire villages.** Vertical dashed lines denote the timing of the four prevalence surveys.

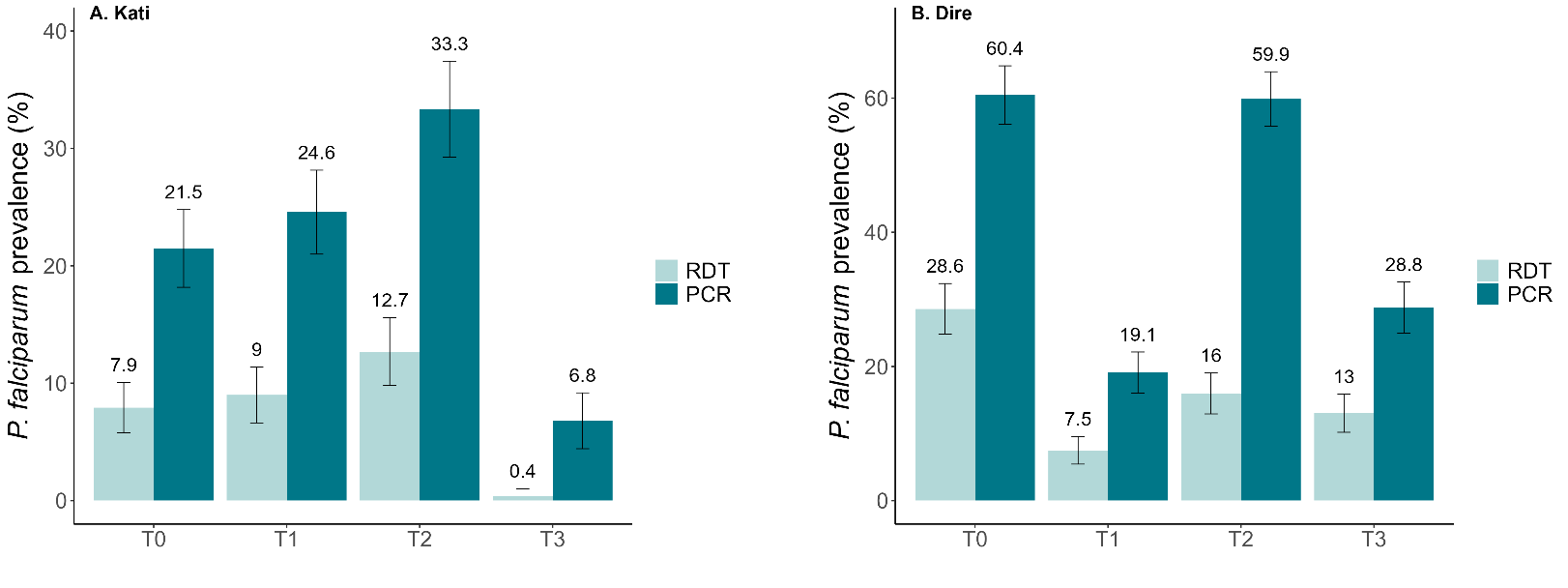

**Figure S6. P. falciparum prevalence among qPCR-positive participants in Kati (A) and Dire (B).** Bars represent 95% confidence intervals.

**Table S2: Rapid Diagnostic Test (RDT) sensitivity (%).** Proportion of qPCR positive infections correctly identified by RDT (95% CI)

A. **Kati**

| Kati T0 | qPCR positive | qPCR negative | Total |
| --- | --- | --- | --- |
| RDT positive | 23 | 2 | 25 |
| RDT negative | 41 | 232 | 273 |
| Total | 64 | 234 | 298 |
| Sensitivity | 35.9% [25.3–48.2] |  |  |
| Kati T1 |  |  |  |
| RDT positive | 25 | 0 | 25 |
| RDT negative | 45 | 216 | 261 |
| Total | 70 | 216 | 286 |
| Sensitivity | 35.7% [25.5–47.4] |  |  |
| Kati T2 |  |  |  |
| RDT positive | 35 | 11 | 46 |
| RDT negative | 57 | 172 | 229 |
| Total | 92 | 183 | 275 |
| Sensitivity | 38.0% [28.8–48.3] |  |  |
| Kati T3 |  |  |  |
| RDT positive | 1 | 0 | 1 |
| RDT negative | 16 | 230 | 246 |
| Total | 17 | 230 | 247 |
| Sensitivity | 5.9% [1.0–27.0] |  |  |

B. **Dire**

| Dire T0 | qPCR positive | qPCR negative | Total |
| --- | --- | --- | --- |
| RDT positive | 77 | 3 | 80 |
| RDT negative | 85 | 102 | 187 |
| Total | 162 | 105 | 267 |
| Sensitivity | 47.5% [40.0–55.2] |  |  |
| Dire T1 |  |  |  |
| RDT positive | 23 | 8 | 31 |
| RDT negative | 36 | 239 | 275 |
| Total | 59 | 247 | 306 |
| Sensitivity | 39.0% [27.6–51.7] |  |  |
| Dire T2 |  |  |  |
| RDT positive | 46 | 2 | 48 |
| RDT negative | 124 | 114 | 238 |
| Total | 170 | 116 | 286 |
| Sensitivity | 27.1% [20.9–34.2] |  |  |
| Dire T3 |  |  |  |
| RDT positive | 37 | 7 | 44 |
| RDT negative | 44 | 196 | 240 |
| Total | 81 | 203 | 284 |
| Sensitivity | 45.7% [35.3–56.5] |  |  |

**Table S3: Kruskal–Wallis results for P. falciparum parasite density differences across age groups by site and survey.**

|  | | A. KATI | | B. DIRE | |
| --- | --- | --- | --- | --- | --- |
|  | Statistic | | *p* value | Statistic | *p* value |
| Visit |  | |  |  |  |
| T0 | 12.6 | | 0.083 | 25.8 | 0.001 |
| T1 | 7.01 | | 0.428 | 7.85 | 0.346 |
| T2 | 7.97 | | 0.335 | 10.1 | 0.182 |
| T3 | 5.94 | | 0.430 | 14.8 | 0.391 |

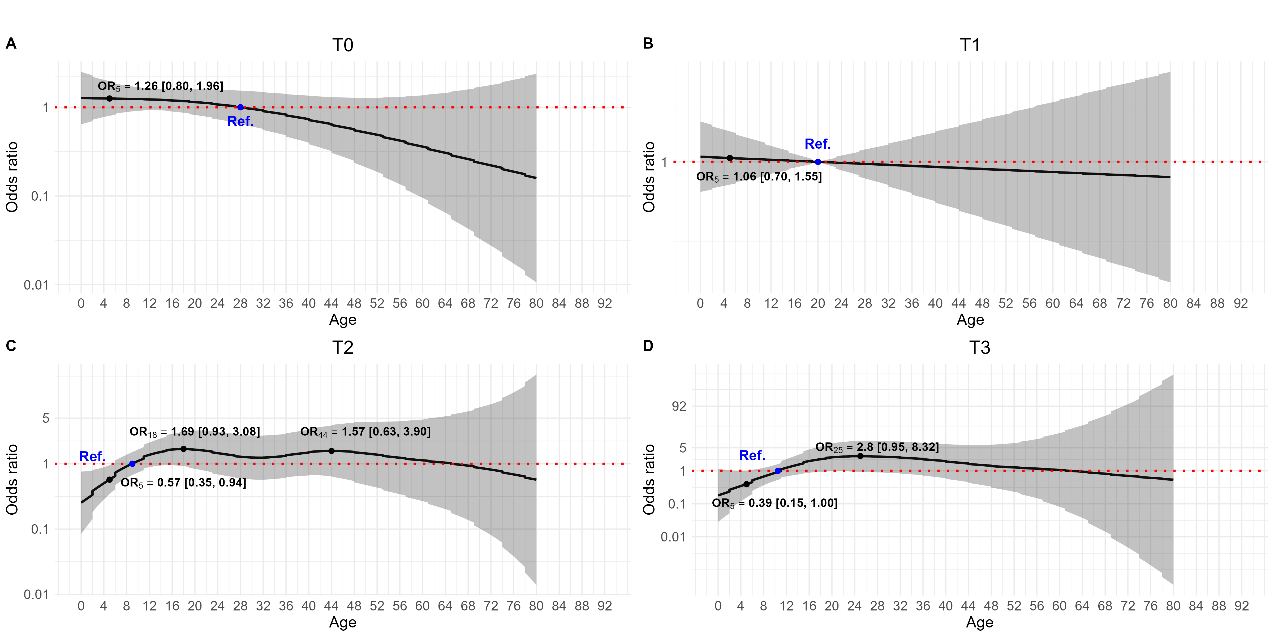

**Figure S7. Multivariate GAMM results linking age to PCR-detected P. falciparum infection across four surveys in Kati.**

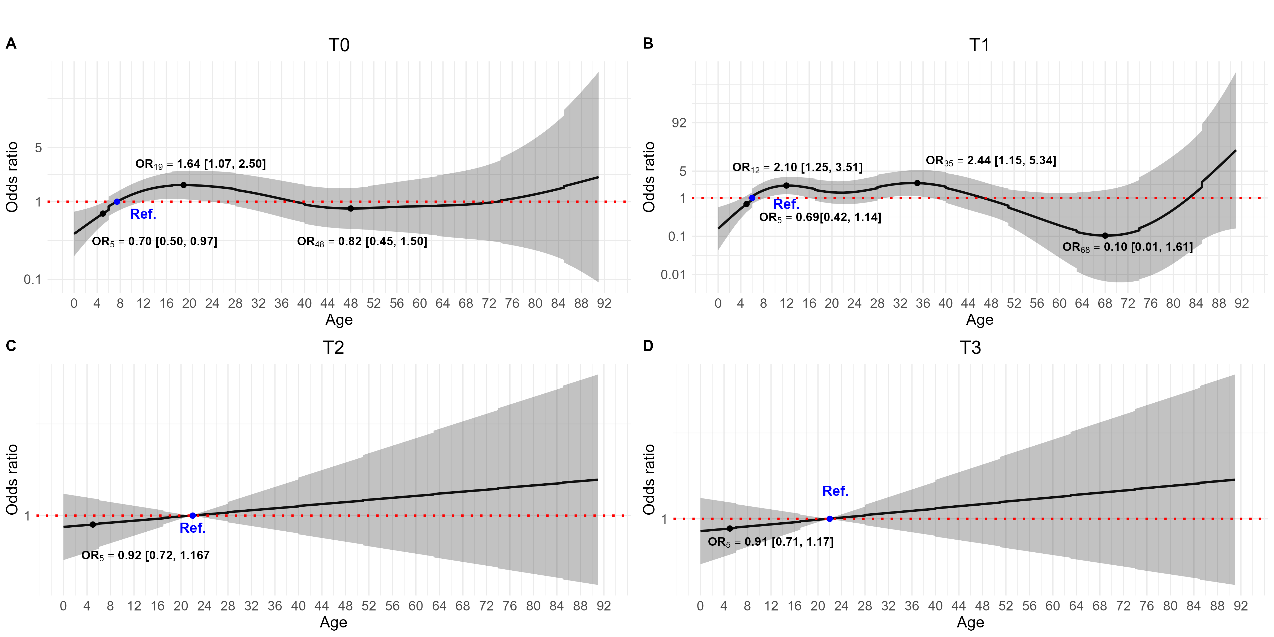

**Figure S8. Multivariate GAMM results linking age to PCR-detected P. falciparum infection across four surveys in Dire.**

**Table S4. Prevention and exposure-related behavioral predictors of P. falciparum infection at T2 from GAMM analyses**

|  | A. KATI | | | | |  | B. DIRE | | | | | | | | | |
| --- | --- | --- | --- | --- | --- | --- | --- | --- | --- | --- | --- | --- | --- | --- | --- | --- |
|  | Univariate analysis | |  | Multivariate analysis | |  | Univariate analysis | | | |  | Multivariate analysis | | | | |
|  | OR [95 CI] | *p* |  | OR [95 CI] | *p* |  | OR [95 CI] | | *p* | |  | OR [95 CI] | | | | *p* |
| SMC during last rainy season |  |  |  |  |  |  |  | |  | |  |  | | | |  |
| Yes | Ref. |  |  |  |  |  |  | |  | |  |  | | | |  |
| No | 9.45 [0.75, 119.02] | 0.0823 |  |  |  |  | |  | |  | | |  |  | | |
| Not eligible >10yrs | 16.29 [3.32, 79.79] | 0.0006 |  |  |  |  | |  | |  | | |  |  | | |
| Bednet use |  |  |  |  |  |  |  | |  | |  |  | | | |  |
| Incorrect use | Ref |  |  | Ref |  |  | Ref | |  | |  | Ref | | | |  |
| Correct use | 0.82 [0.32, 2.12] | 0.6797 |  | 0.83 [0.32, 2.16] | 0.7029 |  | 1.24 [0.61, 2.51] | | 0.5531 | |  | 1.01 [0.43, 2.4] | | | | 0.9811 |
| Outdoor nighttime activity |  |  |  |  |  |  |  | |  | |  |  | | | |  |
| Rarely |  |  |  | Ref |  |  | Ref | |  | |  | Ref | | | |  |
| Often (house activity) | 1.11 [0.49, 2.53] | 0.8054 |  | 1.00 [0.43, 2.31] | 0.9921 |  | 0.69 [0.27, 1.73] | | 0.4245 | |  | 0.59 [0.23, 1.51] | | | | 0.2728 |
| Often (work, vigil) | 1.25 [0.31, 4.95] | 0.7528 |  | 1.2 -[0.3, 4.76] | 0.7953 |  | 0.60 [0.27, 1.33] | | 0.2081 | |  | 0.61 [0.23, 1.59] | | | | 0.3081 |
| Reported mobility |  |  |  |  |  |  |  | |  | |  |  | | | |  |
| No travel | Ref |  |  |  |  |  | Ref. | |  | |  |  | | | |  |
| <1 week within region | 1.34 [0.35, 5.19] | 0.6699 |  |  |  |  | | 0.29 [0.05, 1.58] | | 0.154 | | |  | |  | |
| >1 week within country | 1.40 [0.57, 3.48] | 0.4652 |  |  |  |  | | 0.38 [0.18, 0.80] | | 0.010 | | |  | |  | |
| Unknown | 8.63E+17 | 1 |  |  |  |  | | 2.47 E+28 [0, Inf] | | 1.000 | | |  | |  | |
